# Structural neurodevelopment at the individual level - a life-course investigation using ABCD, IMAGEN and UK Biobank data

**DOI:** 10.1101/2023.09.20.23295841

**Authors:** Runye Shi, Shitong Xiang, Tianye Jia, Trevor W. Robbins, Jujiao Kang, Tobias Banaschewski, Gareth J. Barker, Arun L.W. Bokde, Sylvane Desrivières, Herta Flor, Antoine Grigis, Hugh Garavan, Penny Gowland, Andreas Heinz, Rüdiger Brühl, Jean-Luc Martinot, Marie-Laure Paillère Martinot, Eric Artiges, Frauke Nees, Dimitri Papadopoulos Orfanos, Tomáš Paus, Luise Poustka, Sarah Hohmann, Sabina Millenet, Juliane H. Fröhner, Michael N. Smolka, Nilakshi Vaidya, Henrik Walter, Robert Whelan, Gunter Schumann, Xiaolei Lin, Barbara J. Sahakian, Jianfeng Feng, IMAGEN Consortium

## Abstract

Adolescents exhibit remarkable heterogeneity in the structural architecture of brain development. However, due to the lack of large-scale longitudinal neuroimaging studies, existing research has largely focused on population averages and the neurobiological basis underlying individual heterogeneity remains poorly understood. Using structural magnetic resonance imaging from the IMAGEN cohort (n=1,543), we show that adolescents can be clustered into three groups defined by distinct developmental patterns of whole-brain gray matter volume (GMV). Genetic and epigenetic determinants of group clustering and long-term impacts of neurodevelopment in mid-to-late adulthood were investigated using data from the ABCD, IMAGEN and UK Biobank cohorts. Group 1, characterized by continuously decreasing GMV, showed generally the best neurocognitive performances during adolescence. Compared to Group 1, Group 2 exhibited a slower rate of GMV decrease and worsened neurocognitive development, which was associated with epigenetic changes and greater environmental burden. Further, Group 3 showed increasing GMV and delayed neurocognitive development during adolescence due to a genetic variation, while these disadvantages were attenuated in mid-to-late adulthood. In summary, our study revealed novel clusters of adolescent structural neurodevelopment and suggested that genetically-predicted delayed neurodevelopment has limited long-term effects on mental well-being and socio-economic outcomes later in life. Our results could inform future research on policy interventions aimed at reducing the financial and emotional burden of mental illness.

## Main

Adolescence is a critical and active period for brain reconstruction and maturation, with regional changes of synaptic morphology, dendritic arborization, cortical cell firing and changes in neurochemical receptor affinity ^1–3^. Clinical symptoms of many neuropsychiatric disorders start to emerge during this period, including conduct disorder, mood disorder and schizophrenia^4–7^. Structural neurodevelopment during adolescence is important for enhanced cognitive abilities and mental well-being persisting into adulthood^8–16^. Population-based studies have shown that adolescents exhibit remarkable heterogeneity in terms of structural neurodevelopment^17–19^, but the neurobiological basis of the heterogeneity remains poorly understood. Most efforts have been devoted to study the functional circuitry and structural composition of the brain and their associations with mental health disorders at the population level^1, 20–25^. These pioneering studies have leveraged large population cohorts and refined our understanding of the adolescent brain. However, associations between behavioral patterns and trajectories of brain development vary at the individual level and understanding the sources of variation remains imperative in the arena of public health and precision medicine^12, 18, 26, 27^.

Large-scale longitudinal neuroimaging studies have enabled delineation of the dynamic changes of individual brain morphology, by clustering adolescents according to their developmental trajectories of neuroimaging-derived phenotypes. Similar approaches have yielded associations between atypical brain structure and neuroanatomical variation across neuropsychiatric disorders^28^. Neuroimaging biomarkers offered tremendous versatility to determine the neuropathological mechanisms of neurodegenerative and mental illnesses^29–33^, but have yet not been fully utilized for neurodevelopment. Structural magnetic resonance imaging (sMRI) provides non-invasive measures of imaging-derived phenotypes, among which the developmental courses of gray matter volume (GMV) were shown to be strongly associated with myelinogenesis and synaptic plasticity during adolescence^34–38^. Collectively, this raises the possibility of identifying novel clusters of dynamic brain structure according to the growth trajectories of whole-brain GMV architecture.

In this study, we aim to investigate the individual heterogeneity of adolescent brain development, potential genetic, epigenetic and environmental factors that could contribute to the heterogeneity, and possible long-term impacts of the heterogeneous brain developmental patterns on the biological and social wellbeing later in life. To accomplish these goals, we employ a data-driven approach to cluster adolescents into groups with distinct whole-brain GMV developmental patterns using longitudinal neuroimaging data from the IMAGEN cohort that spanned the entire period of adolescence and early adulthood (schematic workflow in Fig. 1a). Both genome-wide and epigenome-wide association studies are conducted to dissect the genetic and epigenetic changes associated with each cluster.

**Fig. 1.**
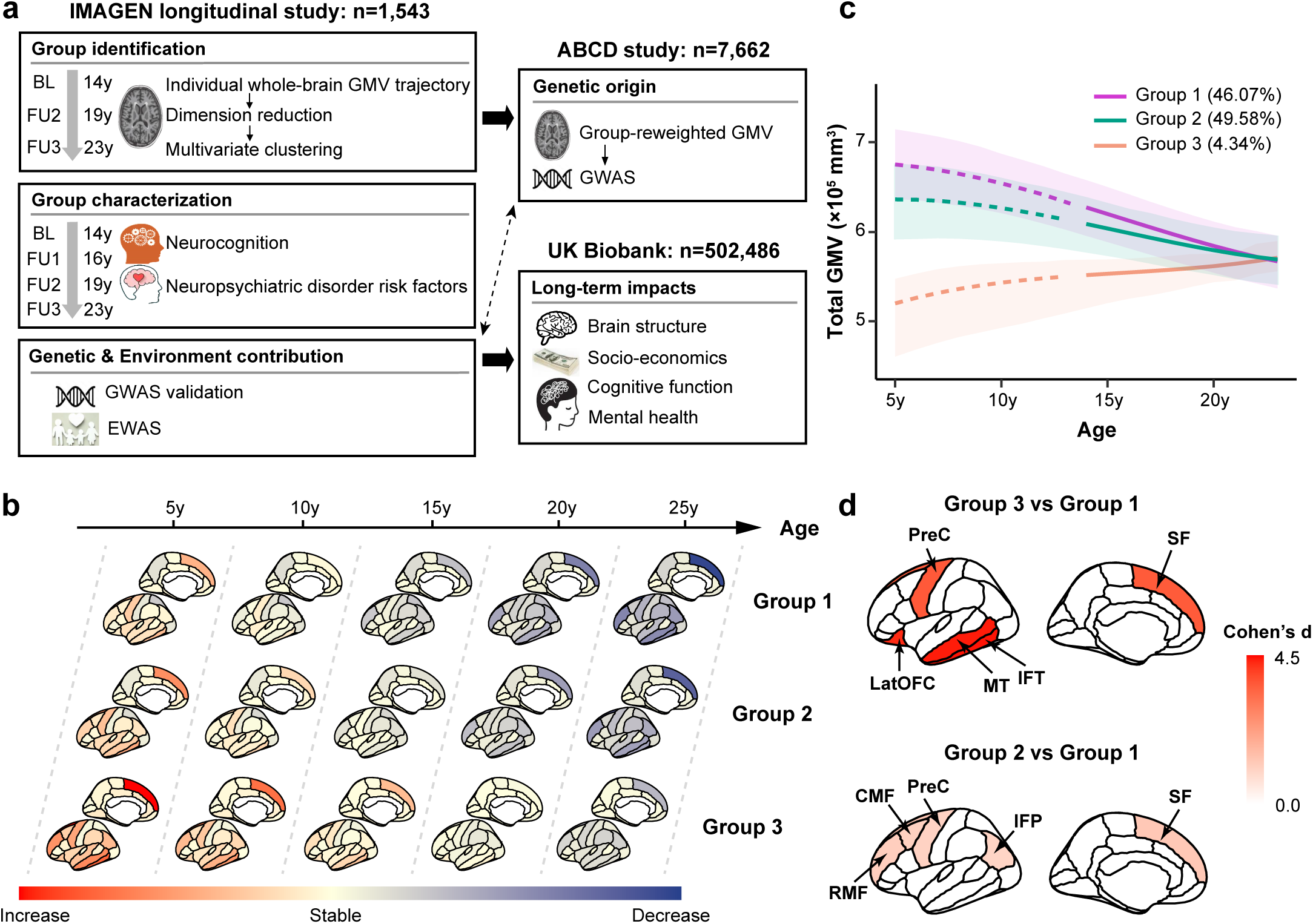
Whole-brain gray matter volume (GMV) developmental patterns define three neurodevelopmental groups. **(a)** Schematic workflow of data sets and analytic methodologies. GMV trajectory in 44 ROIs spanning the cortical and subcortical areas was estimated using linear mixed effect models for adolescents in the IMAGEN study (*n*=1,543). Principal component analysis and multivariate clustering were conducted to identify groups with distinct neurodevelopmental patterns. Next, groups were characterized in terms of neurocognition and risk factors for neuropsychiatric disorders. Genome-wide association study (GWAS) was conducted in the ABCD study (*n*=7,662) using group-reweighted GMV as the proxy phenotype for delayed neurodevelopment, and results were further validated in IMAGEN. Epigenome-wide association study (EWAS) was conducted in IMAGEN (*n*=909) to identify potential environmental exposures associated with neurodevelopment. Last, long-term impacts of the polygenic risk for delayed neurodevelopment on multiple outcomes were investigated among participants in UK Biobank (*n*=502,486) (**Methods**). BL, baseline; FU1, follow-up 1 at 16 y; FU2, follow-up 1 at 19 y; FU3, follow-up 1 at 23 y. **(b)** Whole-brain GMV growth rates (ranging from increase, stable to decrease) at age 5y, 10y, 15y, 20y and 25y were estimated for each group identified in IMAGEN, adjusting for sex, imaging site, handedness and intra-cranial volume (**Methods**). Group 3 showed delayed GMV development compared to Group 1 and 2. **(c)** Total GMV developmental trajectories (with 95% confidence bands) for the three groups identified in the IMAGEN study (Group 1-3). These trajectories were estimated adjusting for sex, imaging site, handedness and intra-cranial volume (**Methods**). Group 1 and 2 exhibited similar GMV developmental trend, while Group 3 had opposite GMV developmental trend. **(d)** The top 5 discriminating ROIs with largest t values comparing the GMV trajectories between Group 3 (*n*=67) and Group 1 (*n*=711) (top), and between Group 2 (*n*=765) and Group 1 (*n*=711) (bottom), adjusting for sex, imaging site, handedness and intra-cranial volume. Two sample t-test: Group 3 vs Group1, IFT (*d*=4.43, *t*=20.13, \*\*\**P_adj_*<0.001), MT (*d*=4.38, *t*=20.07, \*\*\**P_adj_*<0.001), LatOFC (*d*=4.26, *t*=18.31, \*\*\**P_adj_*<0.001), PreC (*d*=3.63, *t*=18.11, \*\*\**P_adj_*<0.001), SF (*d*=3.61, *t*=17.92, \*\*\**P_adj_*<0.001); Group 2 vs Group 1, SF (*d*=1.28, *t*=24.50, \*\*\**P_adj_*<0.001), RMF (*d*=1.14, *t*=21.95, \*\*\**P_adj_*<0.001), CMF (*d*=1.09, *t*=20.77,\*\*\**P_adj_*<0.001), PreC (*d*=1.05, *t*=20.14, \*\*\**P_adj_*<0.001), IFP (*d*=1.00, *t*=19.07, \*\*\**P_adj_*<0.001). LatOFC, lateral orbitofrontal cortex; RMF, rostral middle frontal; CMF, caudal middle frontal; SF, superior frontal; PreC, precentral; MT, middle temporal; IFT, inferior temporal; IFP, inferior parietal.

## Results

### Developmental trajectories of whole-brain GMV during adolescence define three clusters

We began by estimating the longitudinal trajectories of GMV in 44 brain regions of interest (ROIs) (34 cortical and 10 subcortical ROIs) that spanned the whole brain of each adolescent across baseline (at age 14y) and two follow-up scans (at age 19y and 23y) in the IMAGEN study, adjusting for intracranial volume (ICV), sex, handedness and site (Methods). Individuals showed strong heterogeneity and clustering patterns in terms of baseline total GMV and GMV developmental trajectories (Supplementary Fig. 1). Next, we reasoned that neurobiologically meaningful clusters could be explained by the developmental patterns in a subset of ROIs. Therefore, we conducted dimension reduction via principal component analysis (PCA) and selected the first 15 principal components (PCs), which explained 80% of the total variation in whole-brain GMV trajectories, in the clustering analysis (Supplementary Table 1). The first and second PCs defined two combinations of GMV trajectories over the entire brain that were significantly associated with baseline total GMV (Supplementary Fig. 2a). However, they exhibited different association patterns with items of the Cambridge Gambling Task, where PC1 was significantly associated with delay aversion (*r* = 0.07, *P_adj_* = 0.30) and risk adjustment (*r* = - 0.08, *P_adj_* = 0.020), and PC2 was significantly associated with deliberation time (*r* = 0.1, *P_adj_* = 0.003), overall betting (proportion bet) (*r* = 0.07, *P_adj_* = 0.014) and risk-taking (*r* = 0.08, *P_adj_* = 0.008) (Supplementary Fig. 2b). These PCs were then used in the multivariate clustering to identify groups of adolescents with distinct neurodevelopmental patterns.

Among 1,543 adolescents with at least two sMRI scans, our analyses identified three clusters of structural neurodevelopment (𝑃_permutation_ < 0.001) (Supplementary Fig. 3). Group 1 consisted of 711 (46.1%) adolescents, had high baseline total GMV and continuously decreasing GMV at follow-ups, which was consistent with the population GMV developmental trend^28^. Group 2 included 765 (49.6%) adolescents and compared to Group 1, they had lower baseline total GMV, lower peak GMV, and slower rate of GMV decrease. In addition, adolescents in Group 2 are more likely to be older (*Diff* = 0.11y, *P* < 0.001) and be males (*Diff* = 9.5%, *P* < 0.001), have lower parental education (*P* = 0.020 for maternal education; *P*=0.003 for paternal education) and lower WISCIV full score at age 14 (*Diff* = -1.76, *P* = 0.011). The remaining 67 (4.3%) belonged to Group 3, among whom we observed the lowest baseline total GMV and surprisingly increasing GMV at follow-ups, which was opposite to the population developmental trend (Fig. 1b, Supplementary Fig. 4-6). Compared to Group 1, Adolescents in Group 3 are more likely to have lower parental education (*P* = 0.015 for maternal education; *P* = 0.010 for paternal education) and lower WISCIV full score at age 14 (*Diff* = -9.22, *P* < 0.001). The full demographic and baseline characteristics for each group were provided in Supplementary Table 2. Since we aim to investigate group-specific brain developmental patterns in late childhood, we further estimated the age-specific GMV growth rate in each ROI from age 5y to 25y in each group (Methods) using population neurodevelopmental curve as a reference. Consistently we observed continuously decreasing GMV in Group 1 and Group 2 (with slower rate of GMV decrease in Group 2), and increasing GMV in Group 3 for most ROIs (Fig. 1c), indicating delayed neurodevelopment and brain maturation in Group 3 compared to the other groups.

To understand the neurobiological basis of group heterogeneity, we next tested for differences in whole-brain GMV development among these groups. We observed common delayed GMV development in ROIs spanning the inferior temporal, middle temporal, lateral orbitofrontal, precentral and superior frontal areas in Group 3 (relative to Groups 1/2) (Fig. 1d top and Supplementary Table 3). Group 2 showed lower peak GMV and slower rate of GMV decrease in ROIs spanning superior frontal, caudal middle frontal, rostral middle frontal, precentral and inferior parietal areas (relative to Group 1) (Fig. 1d bottom and Supplementary Table 3). These are all among the last areas in the brain to mature and had been implicated to play a key role in executive functions. This led us to ask whether variations in structural neurodevelopment could predict the developmental trajectories of neurocognition and risk of neuropsychiatric disorders in these groups.

### Structural neurodevelopment predicts neurocognition and risk factors for neuropsychiatric disorders

To investigate the association between neurodevelopment and executive functions, we tested for differences of neurocognitive performance among these groups at baseline and at the last follow-up. Full results of these comparisons were provided in Supplementary Table 4. We found that compared to Group 1, Group 3 with delayed neurodevelopment showed worse neurocognitive performance (Spatial Working Memory, Cambridge Gambling Task (CGT) and Stop Signal Task (SST)) at baseline, but most of these items improved over time with brain maturation and became statistically equivalent at the last follow-up (Fig. 2a and Supplementary Fig. 7a). This can be predicted by the structural architecture of GMV development in Group 3, where increasing GMV in the top discriminating ROIs showed positive correlation with improvements of neurocognition (Supplementary Fig. 8 and Supplementary Table 5). In contrast, Group 2 with slower rate of GMV decrease showed worsened neurocognitive performance (CGT and SST) at the last follow-up relative to baseline (Fig. 2a and Supplementary Fig. 7b), which could be predicted by the negative correlations between the GMV developmental trajectories in the top discriminating ROIs and neurocognition (Supplementary Fig. 8 and Supplementary Table 6).

**Fig. 2.**
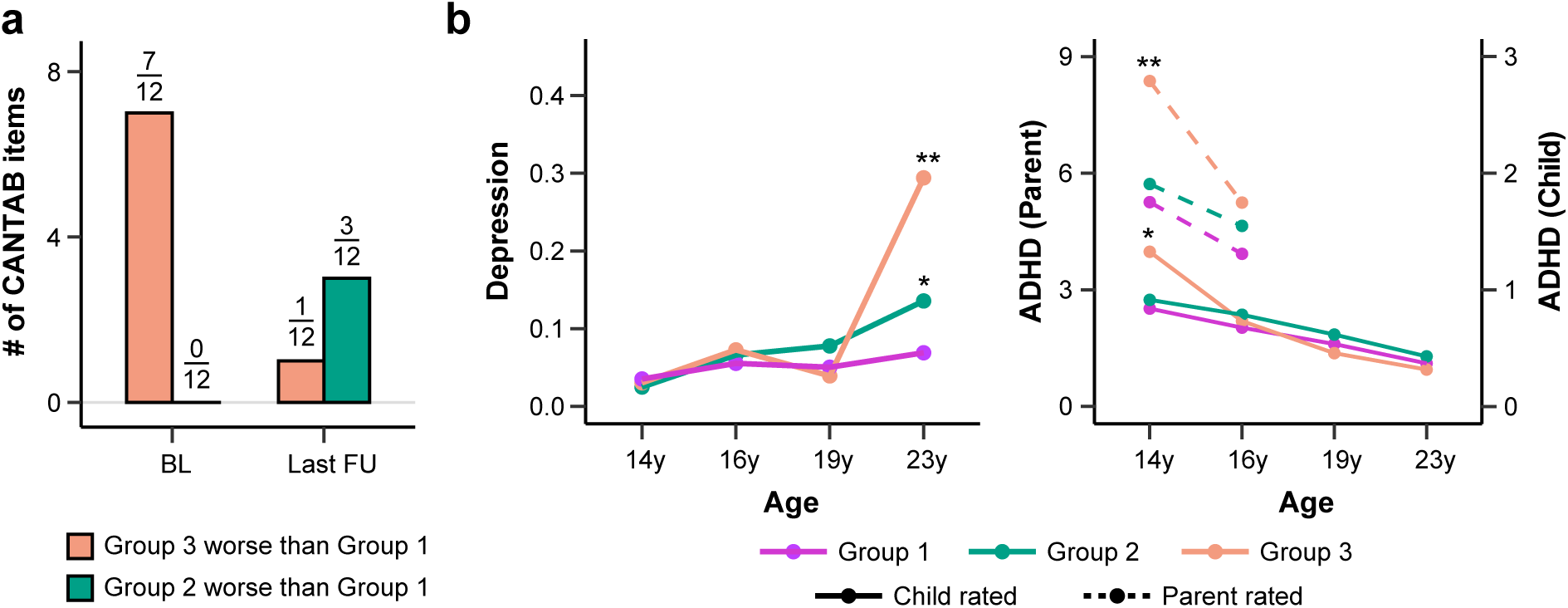
Structural neurodevelopment predicts neurocognition and risk factors for neuropsychiatric disorders. **(a)** Comparison of neurocognitive performances between Group 3 and Group 1 (orange), and between Group 2 and Group 1 (green) at baseline (BL) and the last follow-up (FU) (**Methods**). Total number of neurocognitive tests in cantab where Group 3 performed worse than Group 1 decreased from 7/12 at baseline to 1/12 at follow-up 3, while the number of tests where Group 2 performed worse than Group 1 increased from 0 to 3/12. Full results with item-specific comparisons among these groups are provided in **Supplementary** Fig. 7. CANTAB, Cambridge Neuropsychological Test Automated Battery. **(b)** Longitudinal trajectories of Depression (Left) and ADHD symptoms (Right) among adolescents in the three groups. Group-specific means at each visit were plotted and * indicated significant differences relative to Group 1 adjusting for sex, handedness, stie and ICV. Baseline mental health score was also adjusted for comparison at the last follow-up. Statistical tests were conducted at baseline (14y) and the last follow-up. Depression, Group 2 vs Group 1 at 14y (*d*=-0.05, *P*=0.256), Group 2 vs Group 1 at 23y (*d*=0.13, \**P*=0.023), Group 3 vs Group 1 at 14 y (*d*=-0.01, *P*=0.566), Group 3 vs Group 1 at 23y (*d*=0.70, \*\**P*=0.001); Parent rated ADHD, Group 2 vs Group 1 at 14y (*d*=0.04, *P*=0.220), Group 2 vs Group 1 at 16y (*d*=-0.03, *P*=0.574), Group 3 vs Group 1 at 14 y (*d*=0.34, \*\**P*=0.004), Group 3 vs Group 1 at 16y (*d*=0.01, *P*=0.954); Child rated ADHD, Group 2 vs Group 1 at 14y (*d*=-0.03, *P*=0.321), Group 2 vs Group 1 at 23y (*d*=0.02, *P*=0.758), Group 3 vs Group 1 at 14 y (*d*=0.34, *P*=*0.042), Group 3 vs Group 1 at 23y (*d*=-0.10, *P*=0.579). ADHD, attention-deficit/hyperactivity disorder.

The delayed brain and neurocognitive development in Group 3 led us to ask whether these adolescents had increased risk for neuropsychiatric disorders. Consistent with the improvements of neurocognition, we observed decreased attention-deficit/hyperactivity disorder (ADHD) symptoms, but increased depression symptoms in Group 3 (Fig. 2b and Supplementary Table 4). This indicated that although neurocognitive abilities in Group 3 exhibited pronounced improvement during adolescence, this was not necessarily the case for mental disorder symptoms. Furthermore, consistent with their worsened neurocognitive performances, we observed increased depression symptoms in Group 2 at the last follow-up compared to baseline, (Fig. 2b and Supplementary Table 4). The continuously worsened neurocognition and mental health problems in Group 2 indicated biological, social and mental disadvantages among these adolescents.

Given the slightly different patterns of GMV development for males and females^28^, we conducted the analyses stratified by sex following the same workflow. Results of group clustering largely overlapped with the original analyses (Supplementary Table 7). In general, the sex-stratified analyses revealed similar patterns of neurocognition and mental health symptoms among three groups of adolescents. However, differences of neurocognition among these groups were manifested more for risk-taking and impulsive behaviors in males, while for spatial working memory in females (Supplementary Table 8). Besides, increase of the depressive symptoms in Group 2 was only observed in males, and increase of the depressive symptoms in Group 3 was only observed in females.

In addition, we compared the genetic liability to major neurodevelopmental disorders and related traits, including ADHD, autism spectrum disorder (ASD), educational attainment (EA) and intelligence (IQ), by calculating the corresponding polygenic risk scores (PRS) for each adolescent. Group 3 had higher PRS for ADHD than both Group 1 (*P_adj_* = 0.007) and Group 2 (*P_adj_* = 0.017), while Group 2 was not statistically different from Group 1 (*P_adj_* = 0.42). We did not observe significant differences among the three groups in terms of the PRS of ASD, EA and IQ (Supplementary Table 9). The higher genetic liability of ADHD in Group 3 led us to ask whether genetic variants could explain the delayed neurodevelopment and neurocognitive performances in this group.

### Genetic variation and epigenetic changes contribute to structural neurodevelopment

To better understand the genetic basis of structural neurodevelopment, we conducted genome-wide association studies (GWAS) for Group 3 versus Groups 1/2 using group-reweighted GMV as the proxy-phenotype among 7,662 adolescents in ABCD, since GWAS was under-powered for the IMAGEN study due to limited sample size^39^. Group-reweighted GMV was derived and used as the proxy phenotype because GMV developmental patterns could not be estimated in ABCD due to limited age range. This continuous phenotype represented one’s tendency of being in Group 3 relative to Groups 1/2, or in other words, one’s propensity of having delayed brain development. Specifically, we began by calculating the ROI-specific weight in discriminating Group 3 (relative to Groups 1/2) in IMAGEN adjusting for potential confounders, and applying these weights to corresponding ROIs in ABCD to obtain the Group3-reweighted GMV, which was then used as the proxy phenotype in the Group 3 GWAS (Methods). The Group3-reweighted GMV showed negative correlation with neurocognition in ABCD, indicating the validity of using Group3-reweighted GMV as appropriate proxy for delayed neurodevelopment (Fig. 3a and Supplementary Table 10). Similarly, Group2-reweighted GMV was calculated and used as the proxy phenotype in the Group 2 GWAS.

**Fig. 3.**
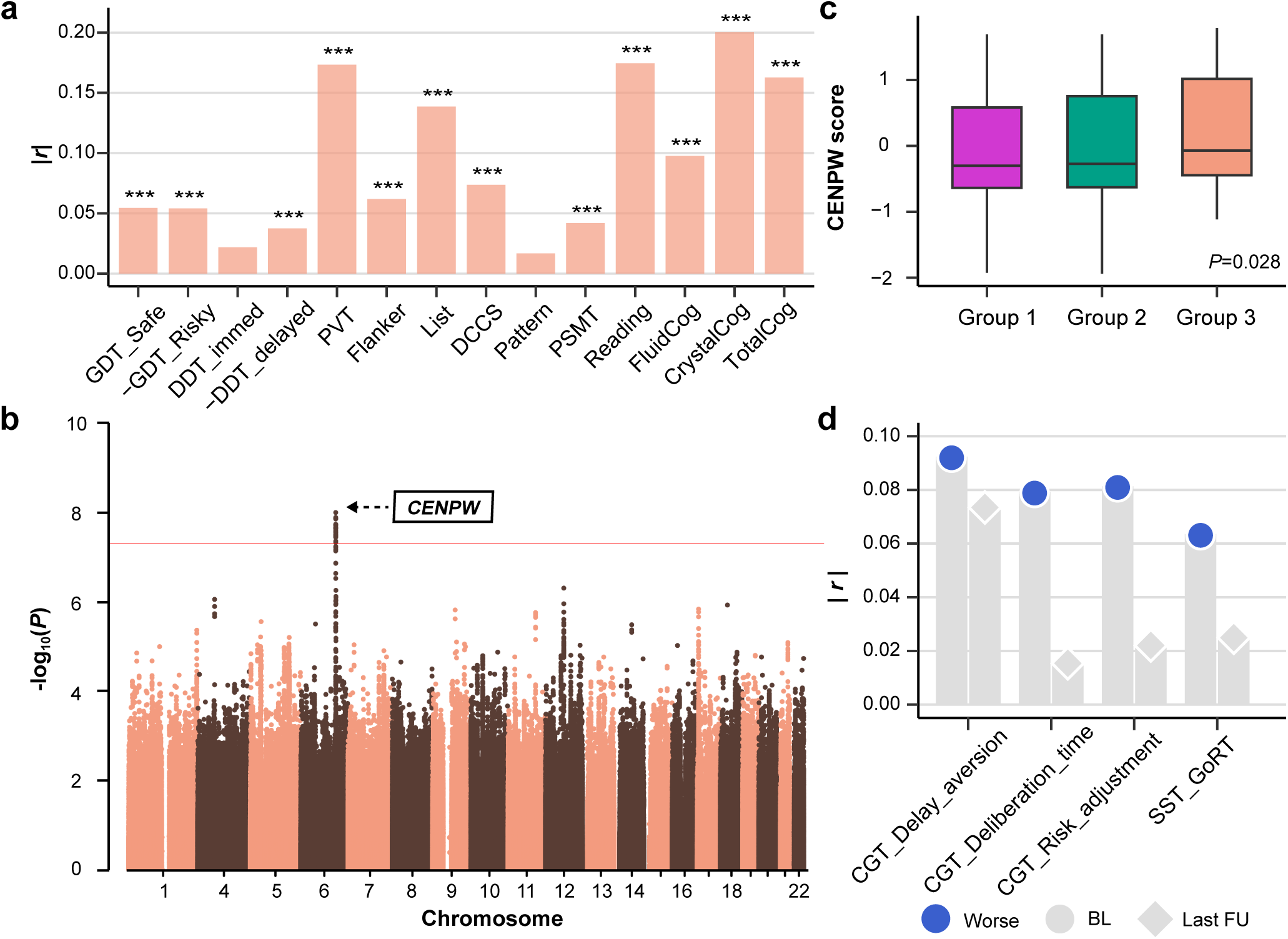
Genome-wide association study (GWAS) identified two significant loci associated with delayed neurodevelopment in Group 3. **(a)** Correlation between Group3-reweighted GMV and neurocognition (**Supplementary Methods**) in ABCD (*n*=11,101) indicated the validity of using the proxy phenotype for delayed neurodevelopment in the following GWAS. GDT, Game of Dice Task; DDT, Dealy Discounting Task; PVT, Picture Vocabulary Test; Flanker, Flanker Inhibitory Control and Attention Test; List, List Sorting Working Memory Test; DCCS, Dimensional Change Card Sort Test; Pattern, Pattern Comparison Processing Speed Test; PSMT, Picture Sequence Memory Test; Reading, Oral Reading Recognition Test; FluidCog, fluid cognition; CrystalCog, crystallized cognition; TotalCog, total cognition. *** *P*<0.001; ** *P*<0.01; * *P*<0.05. **(b)** GWAS Manhattan plot for Group3-reweighted GMV in the ABCD population (*n*=7,662) . Group3-reweighted GMV was calculated for each adolescent (**Methods**) and used as the proxy phenotype for delayed neurodevelopment. Multiple SNPs on chromosome 6 achieved genome-wide significant effects (*P*<5×10^-8^). SNPs on chromosome 6 were mapped to the intronic region of *CENPW*. Results from gene based association analysis (**Supplementary** Fig. 11) confirmed the significant effect of CENPW on delayed neurodeveloopment. Box plot in **(c)** showed that *CENPW* score of delayed neurodevelopment was higher in Group 3 (*n*=60) compared to Group 1 and 2 (*n*=1,338) (two-sided t-test: *P*=0.028). **(d)** indicated that *CENPW* score of delayed neurodevelopment was negatively correlated with baseline (BL) neurocognitive performance, and became non-significant at the last follow-up (FU). Here, *Worse* indicated higher CGT Delay aversion score, lower CGT risk adjustment score, longer CGT Delibration time and SST GoRT. CGT Delay aversion, BL (*r*=0.09, \**P_adj_*=0.027), FU3 (*r*=0.07, *P_adj_*=0.239); CGT Delibration time, BL (*r*=0.08, \**P_adj_*=0.027), FU3 (*r*=-0.02, *P_adj_*=0.983); CGT risk adjustment, BL (*r*=-0.08, \**P_adj_*=0.027), FU3 (*r*=0.022 *P_adj_*=0.983); SST GoRT, BL (*r*=-0.06, \**P_adj_*=0.038), FU3 (*r*=-0.03, *P_adj_*=0.472). CGT, Cambridge Gambling Task; SST GoRT, reaction time for ‘Go’ trials in Stop Signal Task. **(c-d)** confirmed the relationship between *CENPW* and delayed neurodevelopment identified in **(b)**.

One locus showed genome-wide significant effects in the Group 3 GWAS (Fig. 3b and Supplementary Table 11). The lead single-nucleotide polymorphisms (SNP), rs9375442 (*β* = 0.52, *P* = 1.02×10^-8^) on chromosome 6, is an intronic variant located on *CENPW* (Supplementary Fig. 9). *CENPW* is a protein-coding gene involved in the packaging of telomere ends and cell cycle mitotic^40, 41^, and increased *CENPW* expression in progenitors could lead to decreased cortical volume and cognitive function by altering neurogenesis or increasing apoptosis^42^. Other variants on these genes were reported to be associated with cortical surface area and brain volume^43–48^ (Supplementary Fig. 10), general cognitive ability^49–51^ and physical growth^52–56^. Gene-based association analysis confirmed the identification of *CENPW* (Supplementary Fig. 11). Next, we conducted validation of the Group 3 GWAS back in IMAGEN. We began by calculating the polygenic risk score for SNPs (*N_SNP_* = 4) residing in *CENPW* (*CENPW* score) and across the whole genome (PRS) that are associated with Group3-reweighted GMV for each adolescent in IMAGEN, tested for the differences of PRS among these groups, and correlated the PRS with neurocognition and behavioral risk factors. Consistent with the Group 3 GWAS, we observed higher *CENPW* score in Group 3 relative to Groups 1/2 (Fig. 3c) and positive correlations between *CENPW* score and improvement of neurocognition and conduct problems (Fig. 3d). Similar results were obtained for PRS (Supplementary Fig. 12).

No genome-wide significant SNPs were identified in the Group 2 GWAS (Supplementary Fig. 13). However, the large overlap between the neurodevelopmental patterns and homogeneous genetic liability in Group 2 with Group 1 led us to reason that the differences of neurocognitive performances between Group 1 and 2 were quantitative (rather than qualitative) and might subject to the effects of environmental exposure. To test this, we performed epigenome-wide association studies (EWAS) in IMAGEN (Methods) using group label as the phenotype. A significant hypermethylation site cg06064461 (*β* = 25.40, *P* = 4.24×10^-8^) (Fig. 4a) was identified and mapped to *ATF2* and *MIR933* on chromosome 2. *ATF2* encodes a transcription factor of the activator protein-1 family, is ubiquitously expressed in the brain and was found to be associated with both neurodegeneration and neurogenesis^57–59^. *MIR933* shares a common promoter with *ATF2* and offers neuroprotection against neurodegenerative diseases by regulating brain-derived neurotrophic factor (BDNF)^60^. To validate the EWAS results, we correlated the methylation of cg06064461 with estimated GMV trajectory and peak GMV in Groups 1/2, and calculated the mediation effects of cg06064461 methylation in the adverse environment - neurodevelopment pathway. Consistent with the EWAS results, positive correlation between cg06064461 methylation and total GMV trajectory (*r* = 0.14, *P* < 0.001) (Fig. 4b) and negative correlation between cg06064461 methylation and peak GMV (*r* = -0.07, *P* = 0.020) (Fig. 4c) were observed. Although none mediation effects of cg06064461 methylation on the environment – neurodevelopment pathway showed statistical significance after correcting for multiple testing (Fig. 4d and Supplementary Table 12), nominal significance was observed for family affirmation, where higher level of family affirmation was associated with higher peak GMV through reduced cg06064461 methylation (Fig. 4e). Family affirmation was defined as behaviors that the parent puts in place to support or help children in various situations or to show them approval and affection and refers to parent-child relationship^61^. These results indicated that environmental exposure could contribute to disadvantaged neurodevelopment and neurocognition by inducing epigenetic changes of neurogenesis-related genes.

**Fig. 4.**
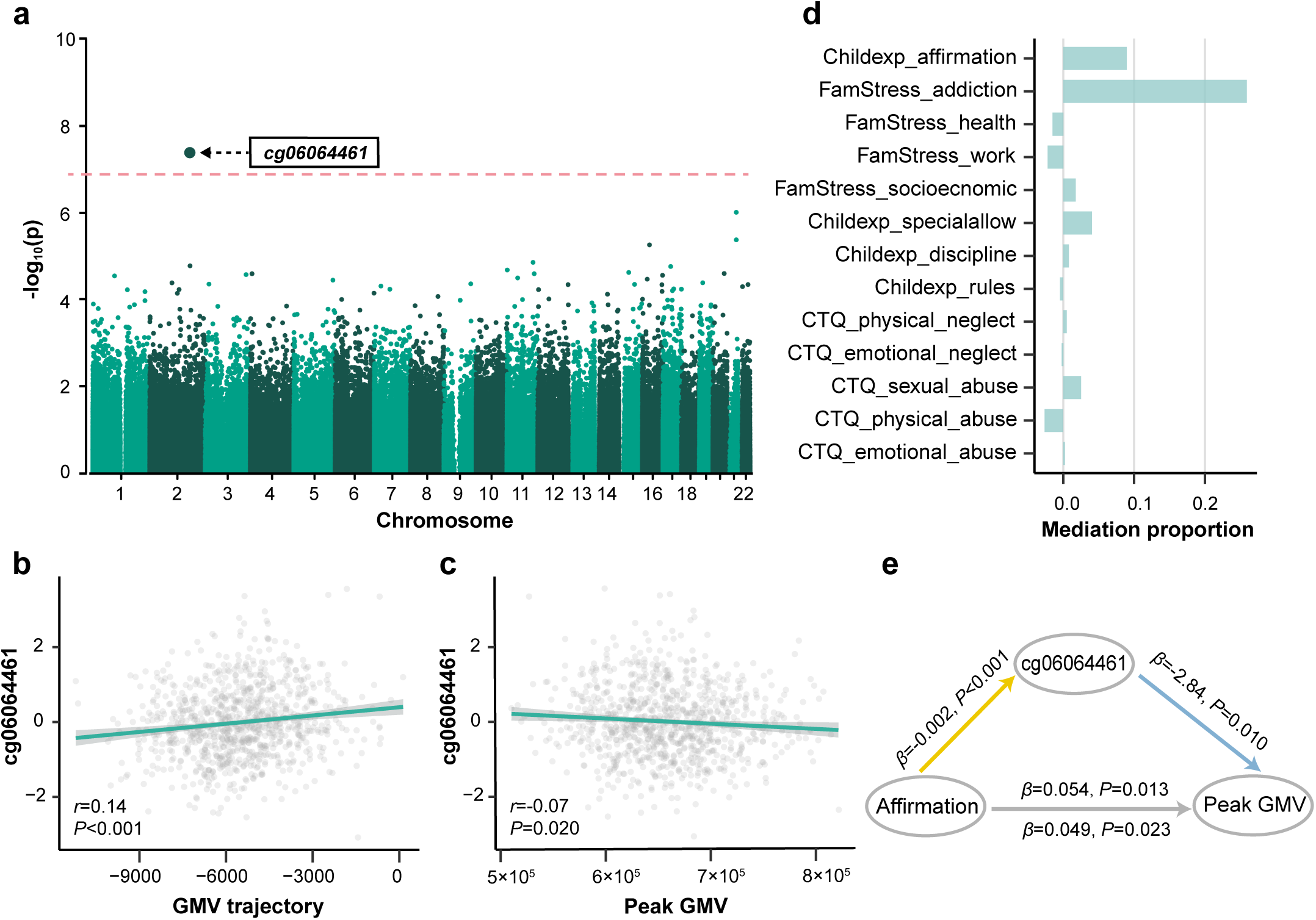
Epigenome-wide association study (EWAS) identified significant signals associated with lowered neurodevelopment in Group 2. **(a)** EWAS Manhattan plot in the IMAGEN population. Group 2 (*n*=463) (relative to Group 1, *n*=446) status was used as the phenotype, adjusting for potential confounders (**Methods**). One hypermethylated site cg06064461 achieved genome-wide significant effect (*P*<5×10^-8^,*P_adj_*<0.05) and was mapped to *ATF2* and *MIR933* on chromosome 2. **(b-c)** Validation of EWAS results in IMAGEN (*n*=909). cg06064461 methylation was positively correlated with total GMV trajectory (**b**; *r*=0.14, \*\*\**P*<0.001) and negatively correlated with peak gray matter volume (GMV) (**c**; *r*=-0.07, \**P*=0.020), adjusting for potential confounders (**Methods**). **(d)** Proportion of the mediation effects through cg06064461 methylation in the environmental exposure - peak GMV pathway, adjusting for potential confounders. Environmental factors were sorted by P values of the mediation effects. Although none mediation effects of cg06064461 methylation showed statistical significance after correcting for multiple testing, nominal significance was observed between family affirmation and peak GMV. Childexp, child’s experience of family life; FamStress, family stressors; CTQ, Childhood Trauma Questionnaire. **(e)** Mediation model was conducted to analyze the direct and indirect effect of family affirmation on peak GMV, with cg06064461 methylation as the mediator. Results showed that cg06064461 methylation significantly mediate the relationship between family affirmation and peak GMV (*β*=0.005, \**P*=0.048).

### Genetic variation had limited effects on the cognitive, mental health and socio-economic outcomes in mid-to-late adulthood

Both genetic vulnerability and structural neurodevelopment are well-established to have profound impact on one’s physical, social and mental well-being in mid-to-late adulthood^62–65^. However, the neurobiological mechanisms through which the long-term effects of genetic variation could be manifested remain largely unknown. We conclude our study by testing in the UK Biobank whether, and to what degree, polygenic risk for delayed neurodevelopment could have impact on the long-term brain structure, cognition, social-economic outcomes and mental well-being.

Motivated by the Group 3 GWAS results, we first calculated the PRS and *CENPW* score of delayed neurodevelopment for each participant in UK Biobank and then correlated them with outcomes of interest. Both PRS and *CENPW* score were approximately normally distributed and negatively associated with total GMV among this population (Fig. 5a). Next, we inspected the association of PRS and *CENPW* score with GMV in multiple brain regions in UK Biobank and found that inferior temporal, fusiform, middle temporal, medial orbitofrontal and lateral orbitofrontal areas were among the most correlated ROIs with PRS of delayed neurodevelopment, and lateral orbitofrontal, caudal middle frontal, rostral middle frontal, insula and superior frontal areas were among the most correlated ROIs with *CENPW* score (Fig. 5b and Supplementary Tables 13-14). These were consistent with the worse spatial working memory among participants with higher PRS of delayed neurodevelopment and *CENPW* score (Supplementary Table 15). Finally, we conducted non-superiority tests of the correlation coefficients and found that correlations between PRS of delayed neurodevelopment and *CENPW* score and all outcomes of interest were smaller than 0.05 (Fig. 5c and Supplementary Figs. 14-15). This indicated that although polygenic risks contributed to delayed neurodevelopment during adolescence, their long-term influences on the cognitive, mental health and socio-economic outcomes were limited once neurocognitive abilities were fully developed.

**Fig. 5.**
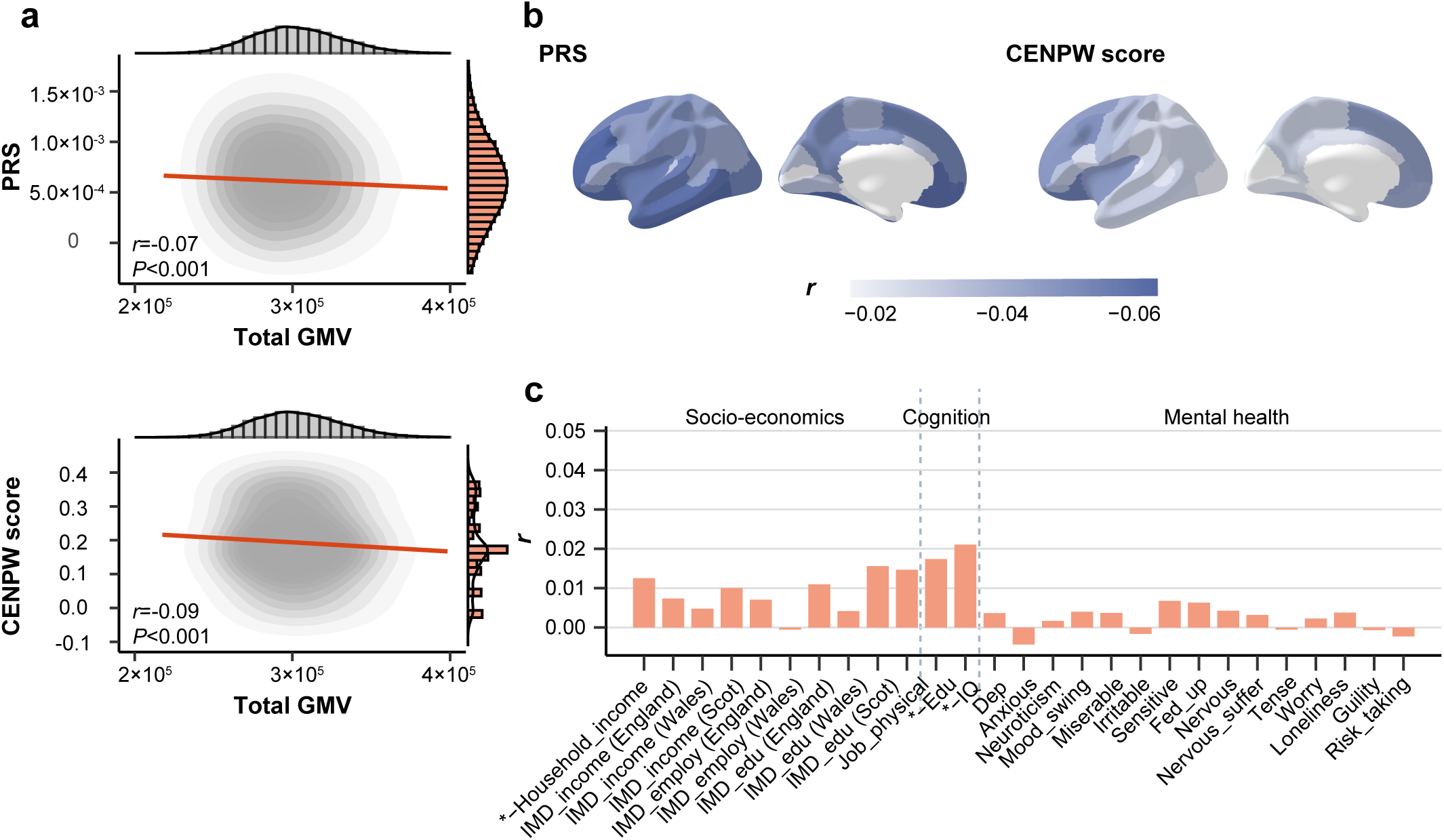
Genetically-predicted neurodevelopment had limited impact on socio-economic, cognitive and mental health outcomes in mid-to-late adulthood. **(a)** Correlation between polygenic risk score (PRS) of delayed neurodevelopment and total gray matter volume (GMV) for participants in UK Biobank (*n*=337,199). Marginal distributions of PRS and total GMV were both normal. PRS and CENPW score both showed negative correlation with total GMV(*r*=-0.07, \*\*\**P*<0.001 for PRS and *r*=-0.07, \*\*\**P*<0.001 for CENPW score). PRS were averaged over different P value thresholds. **(b)** Correlation between averaged PRS of delayed neurodevelopment and *CENPW* score and regional GMV for participants in UK Biobank. Inferior temporal (*r*=-0.07, \*\*\**P_adj_*<0.001), fusiform (*r*=-0.06, \*\*\**P_adj_*<0.001), middle temporal (*r*=-0.06, \*\*\**P_adj_*<0.001), medialorbitofrontal (*r*=-0.06, \*\*\**P_adj_*<0.001) and lateralorbitofrontal (*r*=-0.06, \*\*\**P_adj_*<0.001) were among the ROIs with the strongest correlation with PRS, while lateralorbitofrontal (*r*=-0.06, \*\*\**P_adj_*<0.001), caudalmiddlefrontal (*r*=-0.05, \*\*\**P_ad-j_*<0.001), rostralmiddlefrontal (*r*=-0.05, \*\*\**P_adj_*<0.001), insula (*r*=-0.05, \*\*\**P_adj_*<0.001) and superiorfrontal (*r*=-0.05, \*\*\**P_adj_*<0.001) were ROIs having the strongest correlation with *CENPW* score. These were consistent with the results that participants with higher PRS of delayed neurodevelopment also had worse performance in spatial working memory in UK Biobank. **(c)** Inferiority test of the correlation between averaged PRS and socio-economic, cognitive and mental health outcomes (**Supplementary Methods**) indicated that polygenic risk of delayed neurodevelopment had limited effect on the long-term socio-economic, cognitive and mental health outcomes. Full results were displayed in **Supplementary** Fig. 14. Similar results were observed between CENPW score and these long-term outcomes (**Supplementary** Fig. 15). IMD, Indices of Multiple Deprivation; Scot, Scotland; Edu, the highest educational level; IQ, intelligence.

## Discussion

Adolescence is a dynamic maturational period characterized by potentially suboptimal decision making and an amplified risk of behavioral problems due to the immature brain and cognitive abilities^66–70^. There is a growing consensus that adolescents have remarkably heterogeneous brain developmental patterns^18^. Therefore, studies at the population average level may obscure the true relationship between dynamic brain changes and risks for neuropsychiatric disorders. Here, we developed a data-driven approach that identified three groups of adolescents with distinct whole-brain neurodevelopmental patterns, and showed that these groups had associated genetic or epigenetic determinants, and could predict the paths of both neurocognitive development and long-term socio-economic attainments and mental well-being in mid-to-late adulthood.

Both neuroimaging and animal studies show that gray matter in higher-order brain regions undergoes continuous thinning during adolescence with synaptic pruning and myelination^25, 28, 71, 72^. Therefore, increasing gray matter during this period, especially in higher-order brain regions responsible for executive functions, is indicative of delayed brain maturation. Furthermore, a slower rate of gray matter thinning suggests reduced density of synapses and myelination^3^, which would further limit the enhancement of neurocognitive function and efficient information processing^69, 73, 74^. These diverse growth trajectories of the adolescent brain are capable of shifting both behaviors and the learning capabilities, in ways that could lead to life-long impacts^70, 75–77^. Further, human brain development involves continuing and complex interactions between genetic and environmental influences^23, 26, 78–83^. By integrating genomic, neuroimaging, behavior and health-related data from three large-scale population cohorts, we confirmed that genetic variants are associated with delayed brain maturation and neurocognitive development, without affecting the socio-economic and mental well-being later in life. Whereas, adverse environmental exposure and the associated epigenetic changes could lead to prolonged negative effects on brain development and behavioral disadvantages. Importantly, we regard the differences between Group 2 and Group 1 as quantitative and subject to the magnitude of cumulative adverse environmental exposure, as reflected by the large overlap in their neurodevelopmental patterns and the relatively small effect sizes associated with adverse environmental factors. Consolidating results from EWAS and mediation analysis, our study shed light on the possible epigenetic and neurobiological mechanisms underlying potential causal pathways between environmental exposure and adolescent brain development. However, it does not necessarily mean that the differences between Group 3 and Group 1 could only be attributed to genetic variation, or that differences between Group 2 and Group 1 was purely due to environment. Future research with larger sample size and adequate statistical power are needed to elucidate the potential interplay between gene and environment on structural brain development.

To our knowledge, this is the first attempt to investigate longitudinal brain development at the individual level and its associations with neurocognition and socio-economic outcomes persisting into late adulthood. The three population cohorts involved in our analyses were designed for relatively different purposes, in different populations and produced different data components. Although we tried to link the neurodevelopmental patterns from IMAGEN to ABCD and UK Biobank, this mapping using genetic and neuroimaging associations may subject to confounding bias. For example, the bridging between IMAGEN and ABCD assumed a linear change of GMV from 9 years old (baseline age for the majority participants in ABCD) to 14 years old (baseline age for the majority participants in IMAGEN), and homogenous population composition between these two cohorts. Given the findings from existing investigations (give a reference of brain chart), a linear trend of GMV from 9 to 14 years old were attainable, and in order to achieve population homogeneity, we only selected participants of self-reported “White” ethnicity in ABCD. In addition, both the appropriateness of using the proxy phenotype and results of GWAS conducted in ABCD were successfully validated. However, validation of our results, particularly the long-term impacts in mid-to-late adulthood, are required once long-term follow-ups of socio-economic outcomes in these adolescents become available. Further, the IMAGEN study involves healthy individuals only and our findings may have limited generalizability to specific disease populations. Although these adolescents were not diagnosed with specific neuropsychiatric disorders at baseline, they were likely to be present with subclinical symptoms, referred to minor neurological abnormalities or dysfunction seen in the absence of an obvious cause or pathology. Evidence indicated that subclinical symptoms seen in normal young children were partly attributable to immaturity of the nervous system and were frequently found in the clinical course of psychosis^84^, schizophrenia^85^ and Alzheimer’s disease^86^. Neuroimaging studies thus stand as a powerful tool for identifying important brain regions and morphological phenotypes associated with subclinical symptoms, and for elucidating the neurobiological correlates of subclinical symptoms along the course of brain development. Finally, the three groups identified in this study constitute an initial attempt to solve the problem of heterogeneous brain development that relies heavily on the image-derived phenotypes obtained from sMRI. Further investigation using other neuroimaging modalities, or multi-modal phenotypes are needed for a comprehensive understanding of this dynamic process.

## Methods

### Participants

Genomic, neuroimaging, environmental exposure, behavioral and mental health related data used to identify adolescent neurodevelopmental patterns were obtained from the IMAGEN study. Individuals with GMV beyond 4 interquartile ranges (IQRs) in any ROI were considered as outliers and were excluded from the analyses. After applying the exclusion criteria, 1543 adolescents with at least two structural MRI scans from 14 to 23 years old were included in the analyses (Supplementary Table 16). The average number of structural MRI scans per participant was 2.63, with 974 adolescents having a total of 3 scans (at 14, 19 and 23 years, respectively) and 569 adolescents having a total of 2 scans (384 at 14y and 19y, 147 at 14y and 23y and 38 at 19y and 23y). In addition, genotyping data used in GWAS, validation of GWAS and investigation of the long-term impact were obtained from ABCD, IMAGEN and UK Biobank, respectively. A total of 11,760 participants aged between 9 and 11 years old from ABCD were included, with the average number of structural MRI scans per adolescent 1.68. Further, 502,409 participants aged between 37 and 73 years old from UK Biobank were included in the long-term analyses of structural brain development. Demographics and baseline characteristics of participants from the three large population cohorts were summarized in Supplementary Table 17. A full description of all population cohorts used in the analyses can be found in Supplementary Methods.

## Analysis of Structural MRI data

### Data preprocessing

In brief, quality-controlled processed T1-weighted neuroimaging data were obtained from ABCD, IMAGEN, Human Connectome Project Development (HCP-D), HCP Young Adult (HCP-YA) and the Philadelphia Neurodevelopmental Cohort (PNC). Assessment of regional morphometric structure were extracted by FreeSurfer v6.0 using Desikan-Killiany (h.aparc) atlas for cortical regions, and ASEG atlas for subcortical regions. Quality check was performed according to FreeSurfer reconstruction QC measures. Detailed description of data collection and preprocessing is provided in Supplementary Methods.

### Estimation of GMV developmental trajectory

GMV trajectory in each of the 44 ROIs was estimated for each adolescent using linear mixed effect regression model (*lme4* 1.1-31 package) (since at most three structural MRI scans were available for each adolescent, only random slope model could be robustly estimated). Empirical Bayes estimate of the random slope was extracted for each adolescent. Intracranial volume (ICV), sex, handedness and imaging site were used as covariates to adjust for potential confounding.

### Principal component analysis (PCA) and group clustering

Dimension reduction via PCA was performed to individual GMV trajectories estimated in the 44 ROIs. The first 15 principal components (Supplementary Table 1), which explained 80% of the total variation, were used in the multivariate k-means clustering. The optimal number of clusters was selected based on Elbow method with the constraint that each cluster contain at least 4% of the overall population.

### Permutation test

Permutation was conducted by shuffling the estimated GMV trajectory in each ROI simultaneously and re-performing the dimension reduction and multivariate clustering repeatedly over 1000 times. P value was calculated as the proportion of Between-cluster Sum of Squares / Total Sum of Squares ratio greater than the estimated ratio in the original sample across all 1000 permutations.

### Comparison of GMV trajectory among groups

Pairwise comparisons of GMV developmental trajectories in each ROI among the three groups were conducted via t test. The top 5 ROIs with the largest absolute t values were selected as the top distinguishing ROIs between the corresponding groups (Supplementary Table 3). Cohen’s ds (calculated using *effectsize* 0.8.3 package) for these regions were provided in Fig. 1d and Supplementary Fig. 16.

### Estimation of age and region-specific GMV development among groups

To illustrate the region-specific GMV development in an extended time frame ranging from late childhood to early adulthood, external neuroimaging data from several population cohorts were incorporated. This includes a total of 21,826 participants comprising of 11,811 participants aged 9-14y with 19,587 scans in ABCD, 652 participants aged 5-22y in HCP-D, and 1,587 participants aged 8-23y in PNC study. Since cubic model could not capture GMV trajectory beyond 23y and quadratic model could not utilize data before 9y, we used a reference curve estimated from cross sectional studies (HCP-D + PNC) in estimating the region-specific GMV developmental curve over 5-25y. Distance between GMV in IMAGEN and that in the reference population in the corresponding ROI was used as the dependent variable in the quadratic linear mixed effect model with random intercept and slope, adjusting for ICV and site. Empirical Bayes estimates of the random effects for each group were added to the population averaged estimates to yield the group-specific developmental curve.

### Estimation of group-specific developmental curve of total GMV in IMAGEN

A two-stage estimating procedure was adopted. Optimal model was selected among a series of polynomial mixed effect models using likelihood ratio test. First, population ICV developmental curve over 5-23y was estimated using the above-mentioned population neuroimaging data. Quadratic linear mixed effect models with random intercept at the individual and study level were fitted. To estimate the developmental curve of total GMV in the reference population (ABCD + HCP + PNC), cubic model adjusting for ICV was selected with random intercepts at the individual and study level. To estimate the developmental curve of total GMV in the ABCD and IMAGEN population, cubic model adjusting for ICV was selected with random intercept and slope at the individual level. Empirical Bayes estimates of the random effects were extracted and averaged in each group. Population ICV estimated in stage 1 was used to fit group-specific curves. The 5^th^ and 95^th^ percentile of the group-specific total GMV were calculated as the 95% confidence interval at each age.

### Estimation of peak total GMV in IMAGEN

To estimate the peak total GMV in the IMAGEN population, 1,113 participants aged 22-38y in HCP-YA study were added to the reference population. A similar two-stage estimating procedure was used and the optimal model was selected based on Bayesian information criterion (BIC) and likelihood ratio test. First, population developmental curve of ICV was estimated using mixed effect regression model (*nlme* 3.1-160 package) with random intercept and slope. Basis function involving centered age was determined as the natural spline with 5 degrees of freedom. Interaction effects between age and sex, sex and study were also included in the regression model. Estimated ICV at each age was retained for the following analysis. Next, linear mixed effect regression model with random intercept and slope was fitted for total GMV. Basis function involving centered age was determined to be B spline with 12 degrees of freedom. Interaction effects between age and sex, sex and study were included in the regression model. Peak total GMV was defined as the highest total GMV one can achieve during brain maturation.

### Comparisons of environmental burden, neurocognition, behavior and mental disorder

To assess whether environmental burden, neurocognition, behavioral risk factors and mental symptoms differ by groups, we analyzed their longitudinal measurements at 14y, 16y, 19y and 23y in IMAGEN. Personal traits, including personality, temperament and characters, were obtained from the NEO Five-Factor Inventory (NEO-FFI) and temperament and character inventory (TCI-R). Environmental burden, including prenatal exposures (parental smoking, maternal drinking and maternal medical problems during pregnancy), birthweight, stressful life events, child trauma experiences, child’s experience of family life and family stressors, were obtained from Pregnancy and Birth Questionnaire (PBQ), life-events questionnaire (LEQ), Childhood Trauma Questionnaire (CTQ), and Family Stress Scale and Family Life Questionnaire from development well-being assessment interview (DAWBA). Neurocognitive performances were obtained from Cambridge Neuropsychological Test Automated Battery (CANTAB) tests, Monetary-Choice Questionnaire (KIRBY) and Stop Signal Task (SST) results. Behavioral assessments, including conduct problems and substance use, were obtained from strengths and difficulties questionnaire (SDQ), European school survey project on alcohol and drugs (ESPAD), Fagerstrom test for nicotine dependence (FTND). Mental health conditions, including ADHD and depression, were obtained from self-rated development well-being assessment interview (DAWBA), where Attention-deficit/hyperactivity disorder (ADHD) score was additionally calculated using parent-rated interview. A detailed description of these assessment instruments is provided in Supplementary Methods. Generalized linear models adjusting for sex, handedness and ICV were used for comparing these tests at baseline and at the last follow-up visit (19y for Pattern recognition memory (PRM), Affective Go-No Go (AGN) and Rapid visual information processing (RVP); 23y for all other tests). For child-rated ADHD and depression score, baseline scores were also included as covariates. Intra-Extra Dimensional Set Shift (IED) test was only available at age 23y, and parent-rated ADHD score was only available at 14y and 16y. Cohen’s d was calculated for each measurement after regressing out the covariates. False discovery rate (FDR) was used to correct for multiple testing within scales.

### Quality control of genomic data

In this study, we performed stringent QC standards using PLINK 1.90. Individuals with > 10% missing rate and single-nucleotide polymorphisms (SNPs) with call rates < 95%, minor allele frequency < 0.1%, deviation from the Hardy-Weinberg equilibrium with P < 1E-10 were excluded from the analysis. For ABCD, we only selected subjects with self-reporting White ancestral origins using the public release 3.0 imputed genotype data, which was imputed with the HRC reference panel^87^. Considering that ABCD is oversampled for siblings and twins, we randomly selected one participant within each family. For IMAGEN, details about preprocessing of genomic data can be found in previous reports ^88^ and data was imputed with the HapMap3 reference panel^89^. For UKB, we selected subjects that were estimated to have recent British ancestry and have no more than ten putative third-degree relatives in the kinship table using the sample quality control information provided by UKB. For more details, please refer to the official document for genetic data of the UKB (http://www.ukbiobank.ac.uk/scientists-3/genetic-data/). After quality control, we obtained a total of 4,244,228 SNPs and 7,662 participants in ABCD, 5,966,316 SNPs and 1,982 participants in IMAGEN, and 616,339 SNPs and 337,199 participants in UKB.

### Calculation of genetic liability

For each individual, polygenic risk scores (PRS) of ADHD, ASD, EA and IQ were calculated based on the public GWAS summary statistics^51, 90–92^ using PRSice v2.3.3. For ADHD, ASD and IQ, optimal p-value thresholds were determined based on the best-fit R2 using parent-rated psychiatric scores for ADHD and ASD, and the total WISCIV score. For EA, variants were selected using a P value threshold from 5e-08 to 1 with a step of 5e-05 and an average score under each P value threshold was calculated.

### GWAS and validation

Since it was difficult to estimate individual GMV developmental trajectory in ABCD with limited number of structural MRI scans per participant and limited age range, we calculated the group-reweighted GMV as a proxy phenotype. There are several underlying assumptions in this calculation. Firstly, it assumes that all brain regions exhibit a comparable linear change from childhood to adolescence. Secondly, it assumes that the participants from ABCD and IMAGEN are drawn from a homogeneous population. Once again, we only included individuals in ABCD with self-report White ancestral origins. ROI-specific loading contributing to group classification (Group 2 vs Group 1, Group 3 vs Group 1, Group 3 vs Group 2, and Group 3 vs Groups 1/2) were obtained by regressing baseline GMV in 44 ROIs adjusting for age, sex, handedness and site in IMAGEN. Logistic regression model was used as the classification model and top 10 ROIs with the largest loadings were used to calculate the group-reweighted GMV in ABCD. Since results remained similar when comparing Group 3 vs Group 1 and when comparing Group 3 vs Group 2 (Supplementary Figs. 17-18), we combined Group 1 and 2 for increased statistical power, and performed the GWAS to investigate the genetic variations associated with Group 3 vs Groups 1/2 (delayed brain development). GWAS was conducted in the White population adjusting for sex, scanner effect and top 20 PCA components using Plink 2 (Supplementary Fig. 19). To ensure the validity of group reweighted phenotype, we correlated the Group-3 reweighted GMV and neurocognitive assessments (Game of Dice Task, Delay Discounting Task and NIH Toolbox) in ABCD (Supplementary Methods). Gene-based association analysis was conducted via MAGMA (version 1.10) using raw genomics data with the same covariate adjustment. To validate the GWAS results, polygenic risk scores for SNPs residing in CENPW (referred to as CENPW score) and across the whole genome (referred to as PRS) were calculated. Four SNPs were obtained by clumping within 250 kb upstream and downstream of CENPW (chr6:126339789-126483320) using Plink 2. PRS was calculated using PRSice using the most predictive P threshold for group-reweighted baseline GMV (r=0.06, P=0.033). Distribution of PRS between Group 3 vs Groups 1/2 and correlation coefficients between PRS and neurocognition, behavior and mental disorder at age 14y and 23y were obtained. FDR was used for multiple tests correction within scales.

### EWAS, gene-specific methylation analysis and results validation

EWAS was performed among 909 adolescents in Group 1 and Group 2 in IMAGEN. Methylation data were collected using the Illumina Infinium HumanMethylation450 BeadChip. Locus-specific genome-wise methylation analysis was conducted and beta values at each Autosomal CpG site between groups was compared using logistic regression adjusting for sex, experimental batches (recruitment center and acquisition wave), the first two principal components of methylation composition and the first four principal components of estimated differential cell counts. Statistical significance was set as false discovery rate (FDR) adjusted p-value < 0.05. Next, we aimed to investigate the association between CpG site and gene methylation with environmental factors of interest. We conducted mediation analyses (*sem* function in the *lavaan* 0.6-12 package) and estimated the total effect of childhood experiences on estimated peak GMV and the indirect effect mediated by cg06064461 hypermethylation. Sex, batches effects, methylation composition components and differential cell count components were included as covariates. Total, direct and indirect effect and their standard deviations were estimated using 1000-iterated nonparametric bootstrap approach. False discovery rate (FDR) was used to correct for multiple testing within scales.

### Long-term impacts of neurodevelopment in UK Biobank

Socio-economic, cognitive and mental health outcomes were obtained at baseline visit among participants in UK Biobank. Socioeconomic conditions were assessed by household income, jobs involved in physical activity and Indices of Multiple Deprivation (IMD) (education, employment and income scores). Cognition was assessed by fluid intelligence and the highest educational attainment. The educational level was divided into four ordinal categories: (1) College or University degree; (2) A levels/AS levels, NVQ or HND or HNC, other professional qualifications or equivalent; (3) O levels/GCSEs, CSEs or equivalent; (4) None of the above. Mental health was assessed by diagnosis of mental disorders (anxiety, depression and mania), summary score of neuroticism and self-reported mental symptom appearances. A detailed description of assessment instruments used in the analysis can be found in Supplementary Methods. To estimate the long-term effect of delayed neurodevelopment, we calculated CENPW score and RPS according to the results of Group 3 GWAS and correlated these scores with outcomes of interest after regressing out the age effect at recruitment, site and gender. It should be noted that these scores only reflect a genetic predicted risk for delayed brain development. Given the large age gap between participants in UK Biobank and IMAGEN, it is challenging to disentangle the long-term impacts of neurodevelopment from those due to potential environmental confounding in mid-to-late adulthood. Therefore, this analysis only serves to explore the potential long-term influence of genetically predicted delayed neurodevelopment and does not account for potential confounding due to environmental factors. Similarly, we assume the homogeneity of study populations between IMAGEN and UK Biobank. For PRS calculation, we used P value thresholds from 5E-08 to 1 with a step of 5E-05 and calculated an average PRS score for each individual. Due to the large sample size and easily-obtainable statistical significance, inferiority tests against 0.05 were conducted against the null hypothesis that the absolute correlation coefficient was less than 0.05.

## Data availability

Data from the ABCD study are available from a dedicated database: https://abcdstudy.org/ by applicaton; data from the IMAGEN study are available upon application: https://imagen2.cea.fr; HCP data are available from: https://www.humanconnectome.org/ by applicaton; PNC data are available from dbGaP: https://www.ncbi.nlm.nih.gov/projects/gap/cgi-bin/study.cgi?study_id=phs000607.v3.p2 by application; and UKB data are available from https://biobank.ndph.ox.ac.uk/ by application ID 19542. Summary statistics from published GWAS of ADHD, ASD, EA and IQ are available at https://atlas.ctglab.nl/. Summary statistics of the GWAS for delayed brain development in this study will be uploaded to https://atlas.ctglab.nl/ upon publication.

## Code availability

Primary analyses were conducted in R v4.2.2. Linear mixed effect models were performed using *lme4* 1.1-31 and *nlme* 3.1-160 R packages. Mediation analysis was performed using *lavaan* 0.6-12 R package. PLINK 2.0 was used to perform GWAS and calculate *CENPW* score. MAGMA v1.10 was used to perform the gene-based association analysis. PRSice v2.3.3 was used to calculate the PRS. Custom code that supports the main findings of this study will be available https://github.com/abnmsry. Additional information related to this paper are available from the authors upon reasonable request.

## Acknowledgments and funding

This work received support from the following sources: National Key R&D Program of China (No.2019YFA0709502), National Key R&D Program of China (No.2018YFC1312904), Shanghai Municipal Science and Technology Major Project (No.2018SHZDZX01), ZJ Lab, and Shanghai Center for Brain Science and Brain-Inspired Technology, the 111 Project (No.B18015), the European Union-funded FP6 Integrated Project IMAGEN (Reinforcement-related behaviour in normal brain function and psychopathology) (LSHM-CT-2007-037286), the Horizon 2020 funded ERC Advanced Grant ‘STRATIFY’ (Brain network based stratification of reinforcement-related disorders) (695313), Human Brain Project (HBP SGA 2, 785907, and HBP SGA 3, 945539), the Medical Research Council Grant ’c-VEDA’ (Consortium on Vulnerability to Externalizing Disorders and Addictions) (MR/N000390/1), the National Institute of Health (NIH) (R01DA049238, A decentralized macro and micro gene-by-environment interaction analysis of substance use behavior and its brain biomarkers), the National Institute for Health Research (NIHR) Biomedical Research Centre at South London and Maudsley NHS Foundation Trust and King’s College London, the Bundesministeriumfür Bildung und Forschung (BMBF grants 01GS08152; 01EV0711; Forschungsnetz AERIAL 01EE1406A, 01EE1406B; Forschungsnetz IMAC-Mind 01GL1745B), the Deutsche Forschungsgemeinschaft (DFG grants SM 80/7-2, SFB 940, TRR 265, NE 1383/14-1), the Medical Research Foundation and Medical Research Council (grants MR/R00465X/1 and MR/S020306/1), the National Institutes of Health (NIH) funded ENIGMA (grants 5U54EB020403-05 and 1R56AG058854-01), NSFC grant 82150710554 and environMENTAL grant. Further support was provided by grants from: - the ANR (ANR-12- SAMA-0004, AAPG2019 - GeBra), the Eranet Neuron (AF12-NEUR0008-01 - WM2NA; and ANR-18-NEUR00002-01 - ADORe), the Fondation de France (00081242), the Fondation pour la Recherche Médicale (DPA20140629802), the Mission Interministérielle de Lutte-contre-les-Drogues-et-les-Conduites-Addictives (MILDECA), the Assistance-Publique-Hôpitaux-de-Paris and INSERM (interface grant), Paris Sud University IDEX 2012, the Fondation de l’Avenir (grant AP-RM-17-013), the Fédération pour la Recherche sur le Cerveau; the National Institutes of Health, Science Foundation Ireland (16/ERCD/3797), U.S.A. (Axon, Testosterone and Mental Health during Adolescence; RO1 MH085772-01A1) and by NIH Consortium grant U54 EB020403, supported by a cross-NIH alliance that funds Big Data to Knowledge Centres of Excellence. The funders had no role in study design, data collection and analysis, decision to publish or preparation of the manuscript.

