## Supplementary Figures for "Structural neurodevelopment at the individual level - a life-course investigation using ABCD, IMAGEN and UK Biobank data"

### Contents of Supplementary Figures

Supplementary Fig. 1. The joint distribution of baseline total GMV and GMV developmental trajectories ······ 3

Supplementary Fig. 2. The first two PCA components ······ 4

Supplementary Fig. 3. The Elbow Method showing the optimal k ······ 5

Supplementary Fig. 4. Total GMV developmental curves of groups from 14y to 23y ······ 6

Supplementary Fig. 5. Estimated total GMV developmental curves of individuals in groups from 5y to 37y ······ 7

Supplementary Fig. 6. Estimated total GMV developmental curves of groups from 5y to 37y ······ 8

Supplementary Fig. 7. Neurocognition comparisons of Group 3 and Group 2 vs Group 1 at baseline and last follow-up ······ 9

Supplementary Fig. 8. Correlation between longitudinal trajectory of GMV in each cortical ROI and neurocognition in groups ······ 10

Supplementary Fig. 9. Locus zoom of the top SNP rs9375442 on chromosome 6 ······ 11

Supplementary Fig. 10. Associations between SNPs on CENPW and cortical surface area (CSA) and brain volume (BV) in existing studies12

Supplementary Fig. 11. Gene-based GWAS results for delayed neurodevelopment ······ 14

Supplementary Fig. 12. Validation of GWAS results for delayed

neurodevelopment in IMAGEN ··············································· 15
Supplementary Fig. 13. GWAS results and validation for Group 2 vs Group 1 ······································································· 16
Supplementary Fig. 14. Correlations between all PRS and socio-economics, cognitive and mental health and in UKB ····················· 17
Supplementary Fig. 15. Correlations between CENPW score and socio-economics, cognitive and mental health and in UKB ····················· 18
Supplementary Fig. 16. The top 5 discriminating ROIs with largest t values comparing the GMV trajectories between Group 3 and Group 2 ········································································ 19
Supplementary Fig. 17. Miami plot of GWAS for Group 3 vs Group 1 and Group 2 ······································································· 20
Supplementary Fig. 18. Q-Q plots of GWAS for Group 3 vs Group 1 and Group 2 ······································································· 21
Supplementary Fig. 19. Q-Q plot of GWAS for Group 3 vs Group 1/2 ········································································ 22

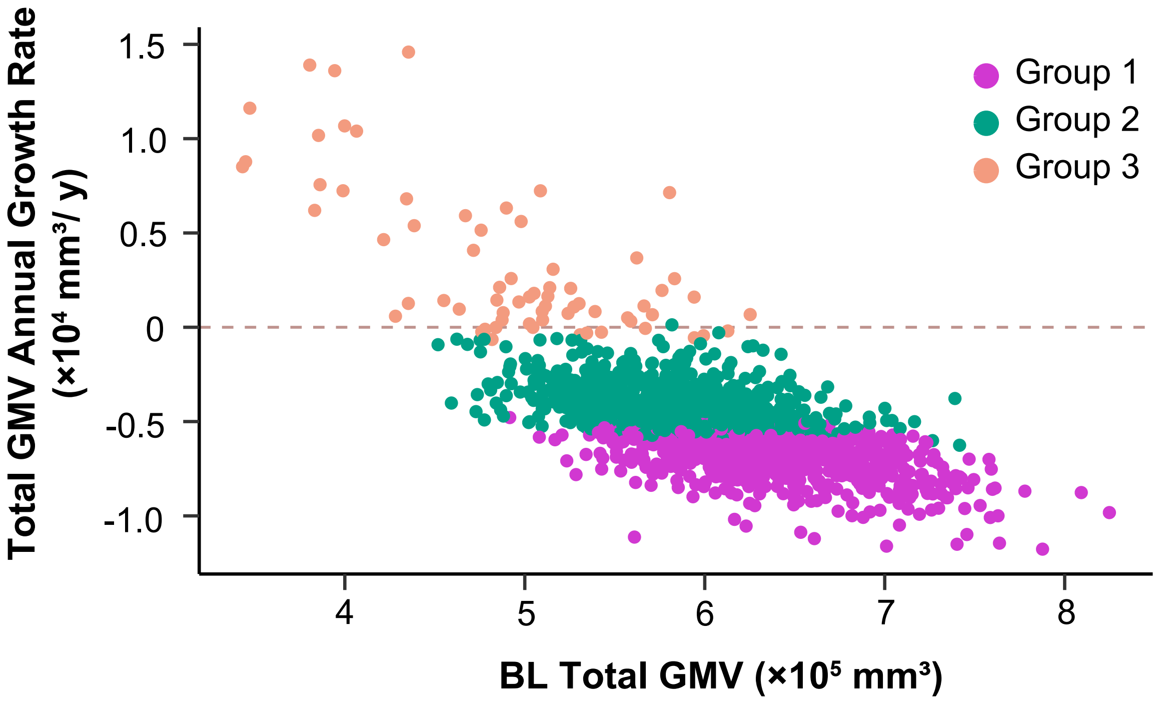


**Supplementary Fig. 1. The joint distribution of baseline total GMV and GMV developmental trajectories.** Individuals in IMAGEN (*n*=1,543) exhibit remarkable heterogeneity in baseline (BL) total GMV and annual growth rate of total GMV from baseline to follow-ups.

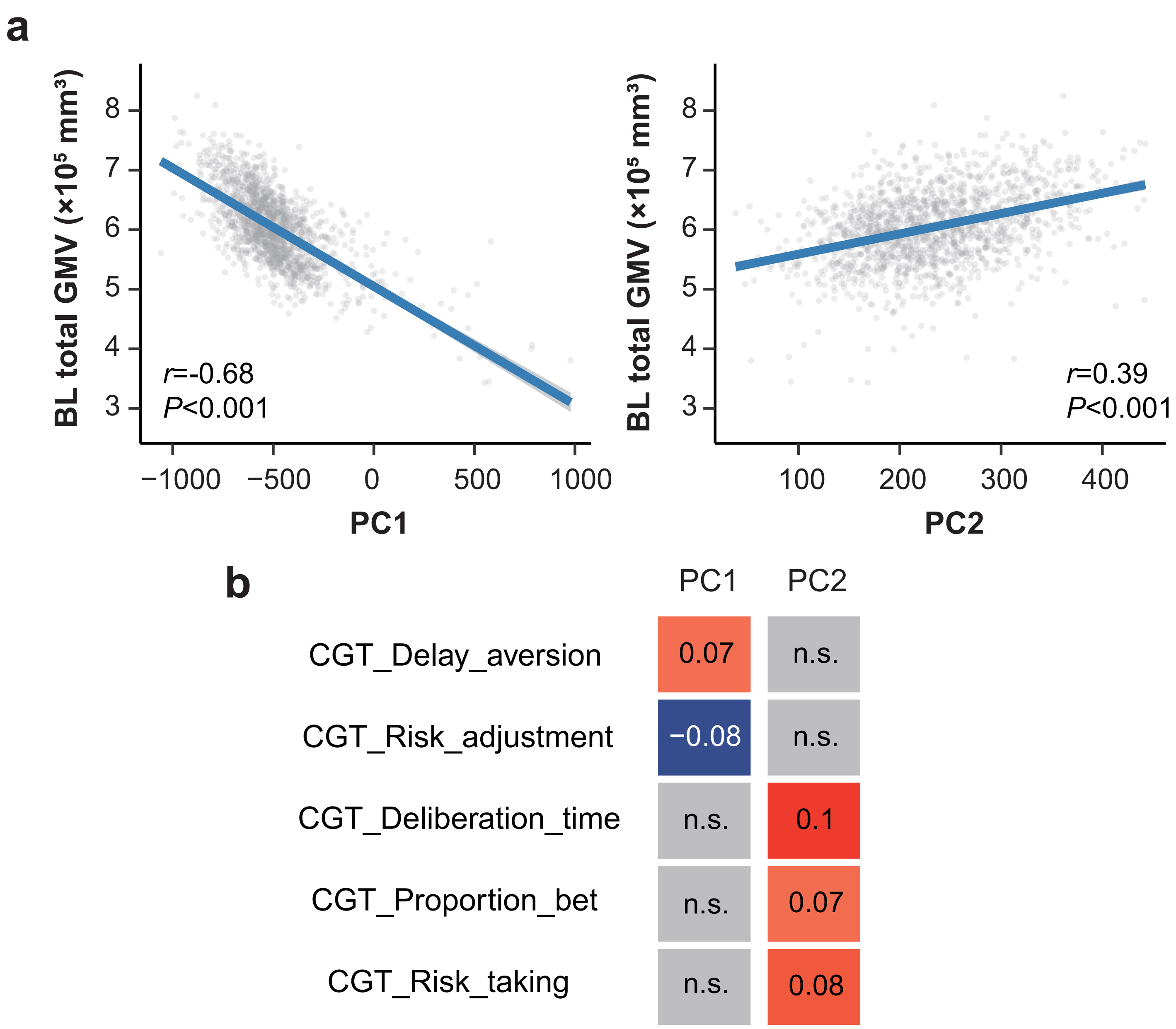


**Supplementary Fig. 2. The first two PCA components.** Principal component analysis was used to define a low-dimensional representation of GMV developmental patterns. The first and second PCs were significantly associated with baseline total GMV **(a)** (*n*=1,543) while they exhibited different association patterns with executive functions in the Cambridge Gambling Task (CGT) **(b)**. **(b)** illustrates the correlations between PCs and CGT performances (n.s. = not significant).

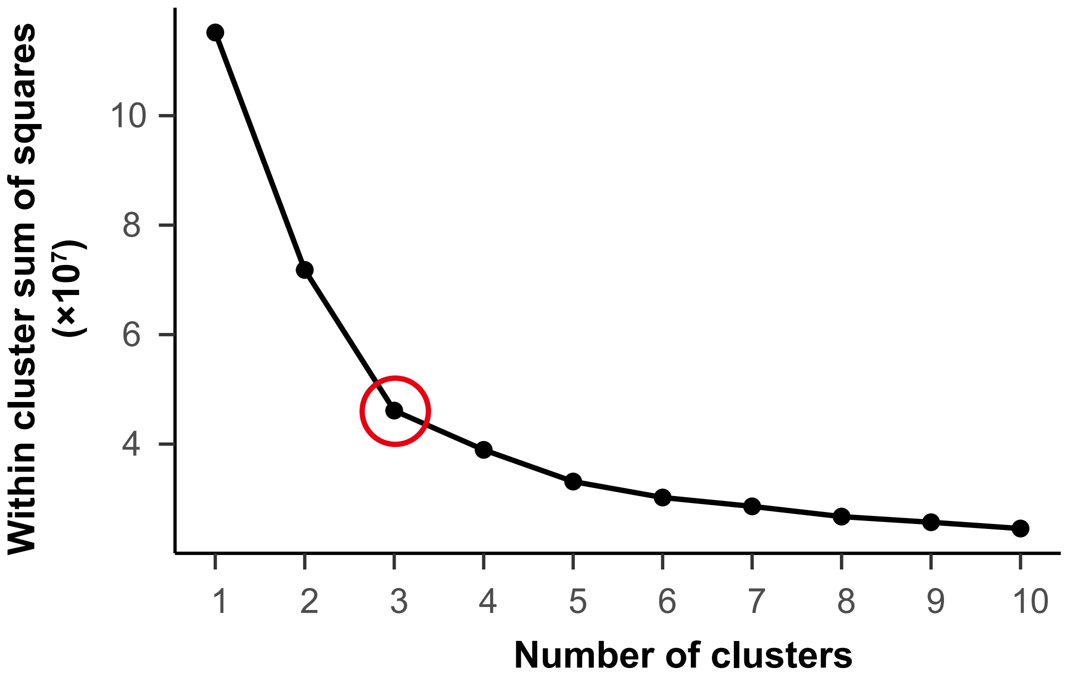


**Supplementary Fig. 3. The Elbow Method showing the optimal k.** Considering the distribution of population among groups, 3 was chosen as the optimal number of cluster.

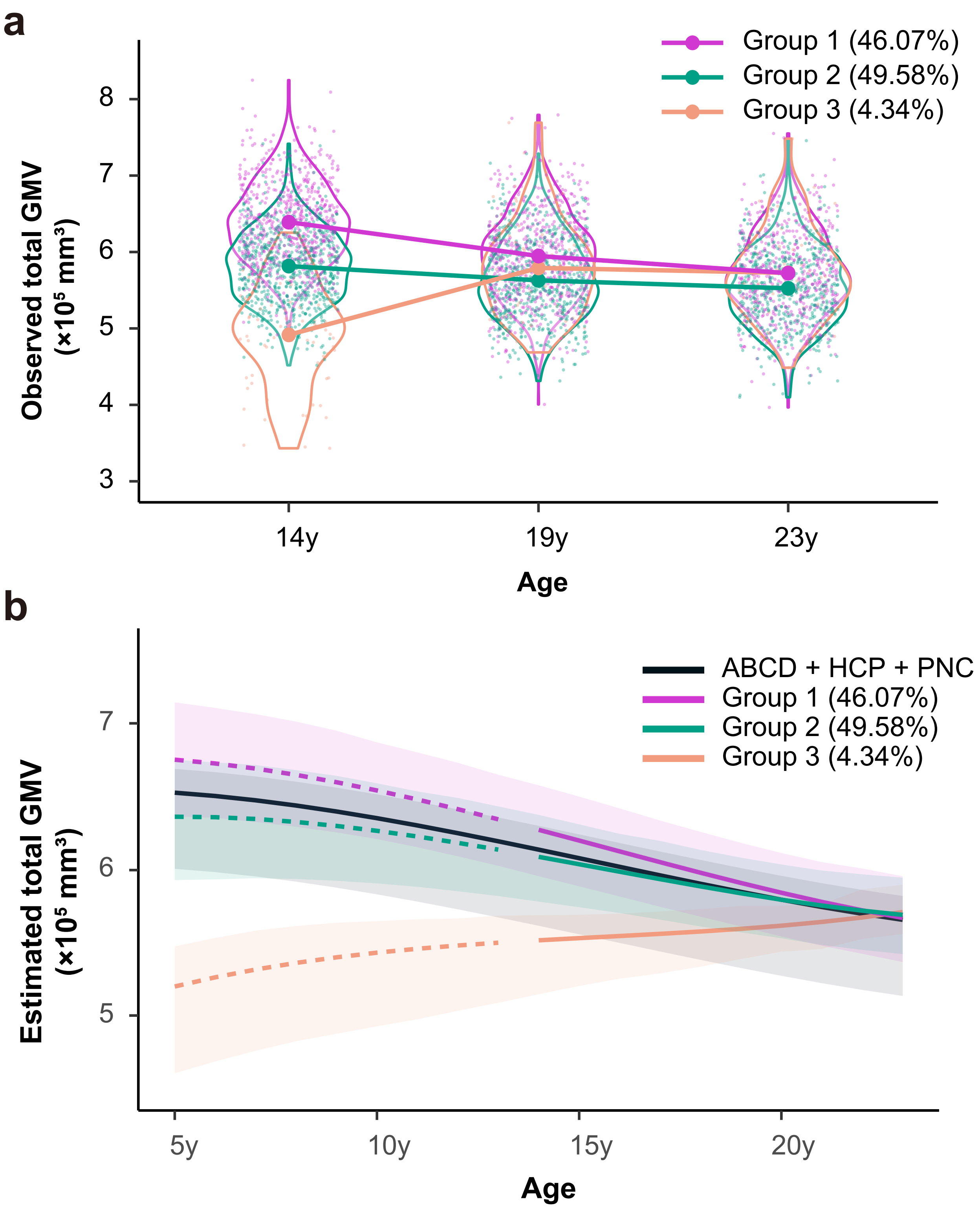


**Supplementary Fig. 4. Total GMV developmental curves of groups from 14y to 23y. (a)** Observed total GMV developmental curves of groups from 14y to 23y (*n*=711 for Group 1, *n*=765 for Group 2, *n*=67 for Group 3). **(b)** Estimated total GMV developmental curves (with 95% confidence bands) for groups and reference population (ABCD+HCP+PNC, *n*=21,826). These curves were estimated adjusting for sex, site/scanner, handedness and intra-cranial volume (**Methods**). Group 1 and 2 exhibited similar GMV developmental trend with the reference population, while Group 3 had opposite GMV developmental trend.

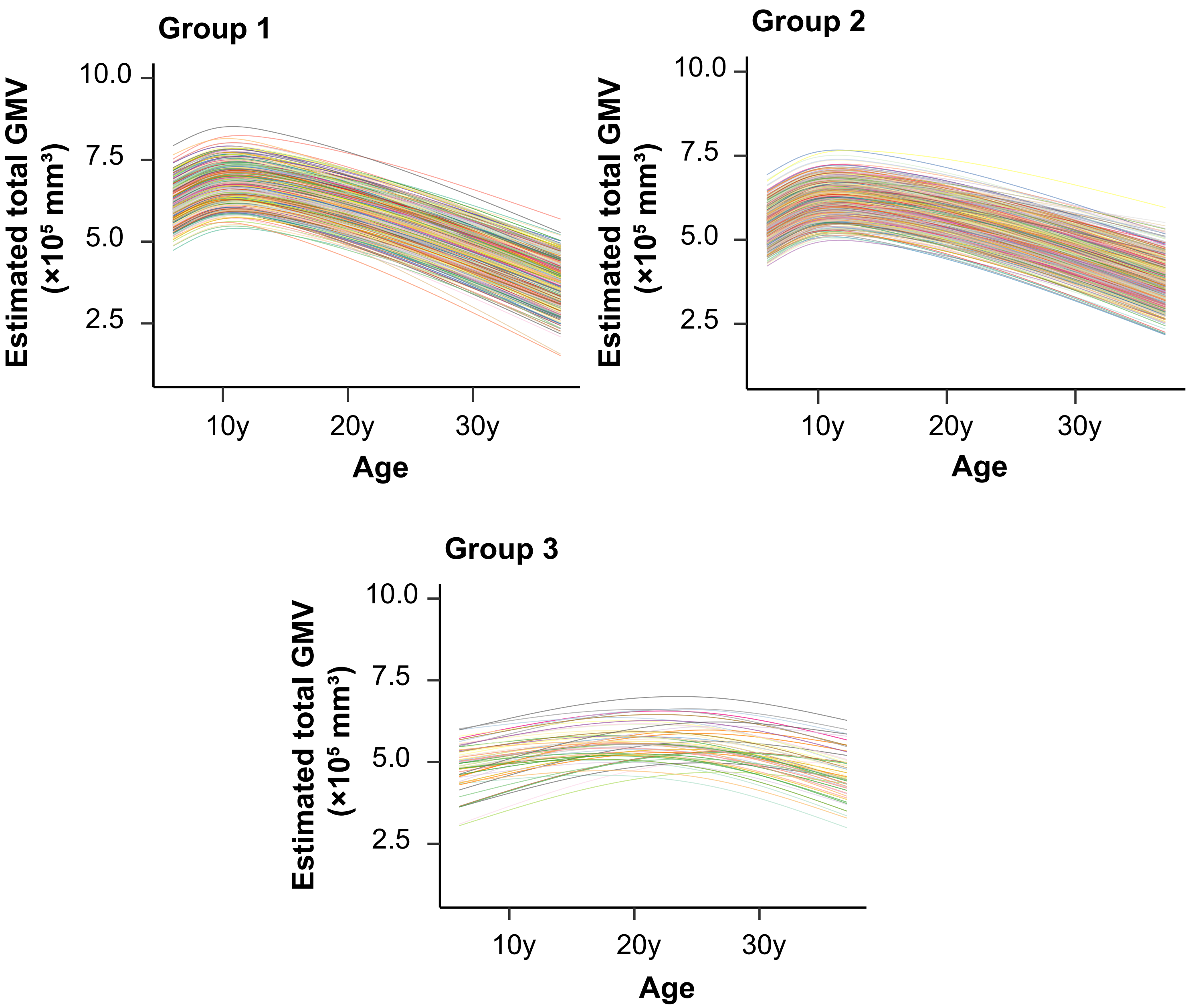


**Supplementary Fig. 5. Estimated total GMV developmental curves of individuals in groups from 5y to 37y.** Total GMV developmental trajectories for individuals in three groups were estimated using linear mixed effect model with B spline function of age (**Methods**).

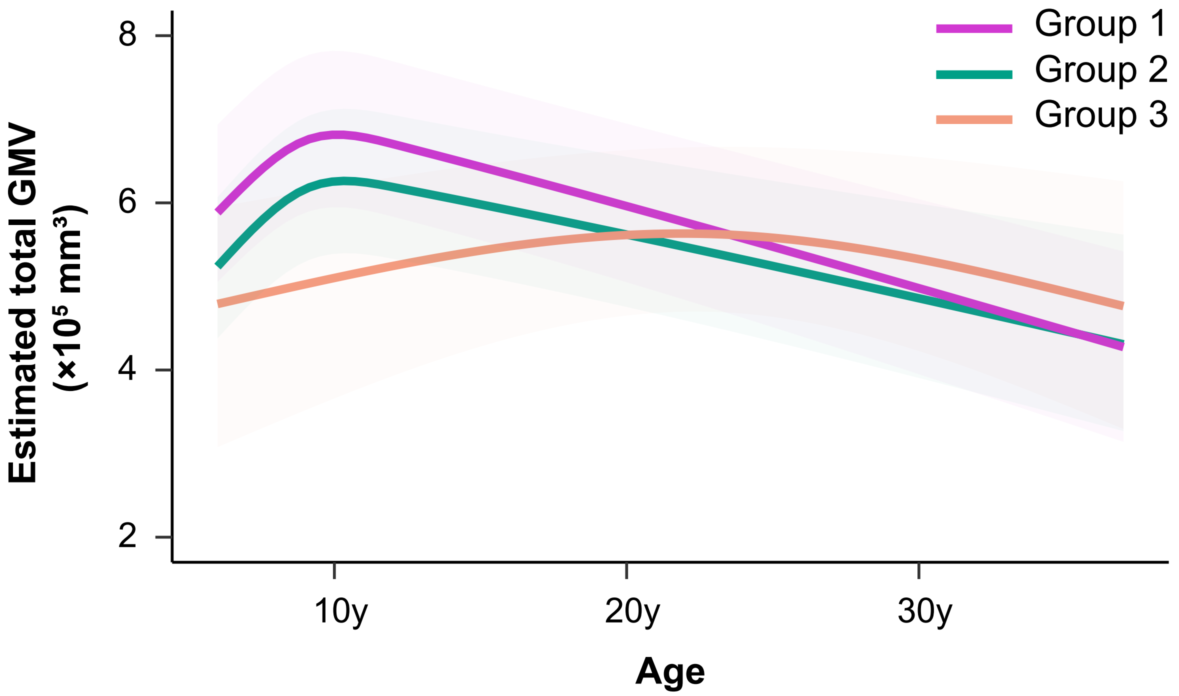


**Supplementary Fig. 6. Estimated total GMV developmental curves of groups from 5y to 37y.** Mean total GMV developmental trajectories (with 95% confidence bands) for groups were plotted using estimated individual GMV trajectories in **Supplementary Fig. 5**. Ranges from the 2.5th percentile to the 97.5th percentile of the corresponding group were plotted as bands.

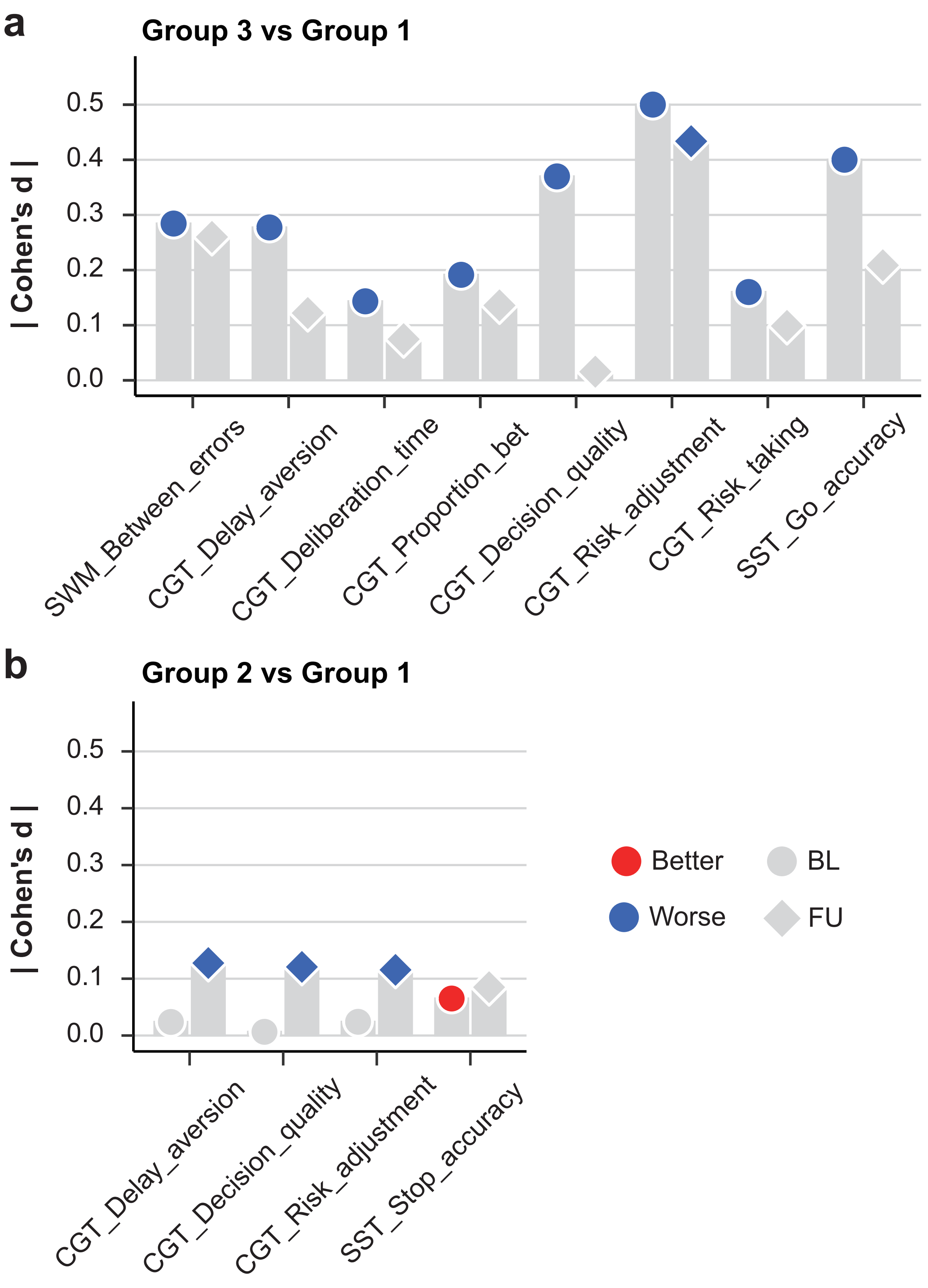


**eSupplementary Fig. 7. Neurocognition comparisons of Group 3 and Group 2 vs Group 1 at baseline and last follow-up.** Neurocognition profiles of Group 3 **(a)** and Group 2 **(b)** at baseline (14y) and last follow-up (23y). Comparisons of neurocognitive performances were converted to cohen's d and item-specific neurocognitive comparisons reflect better or worse performances relative to Group 1. Gray color indicated not significant results. SST comparisons were not included in the count in **Fig. 2a**.

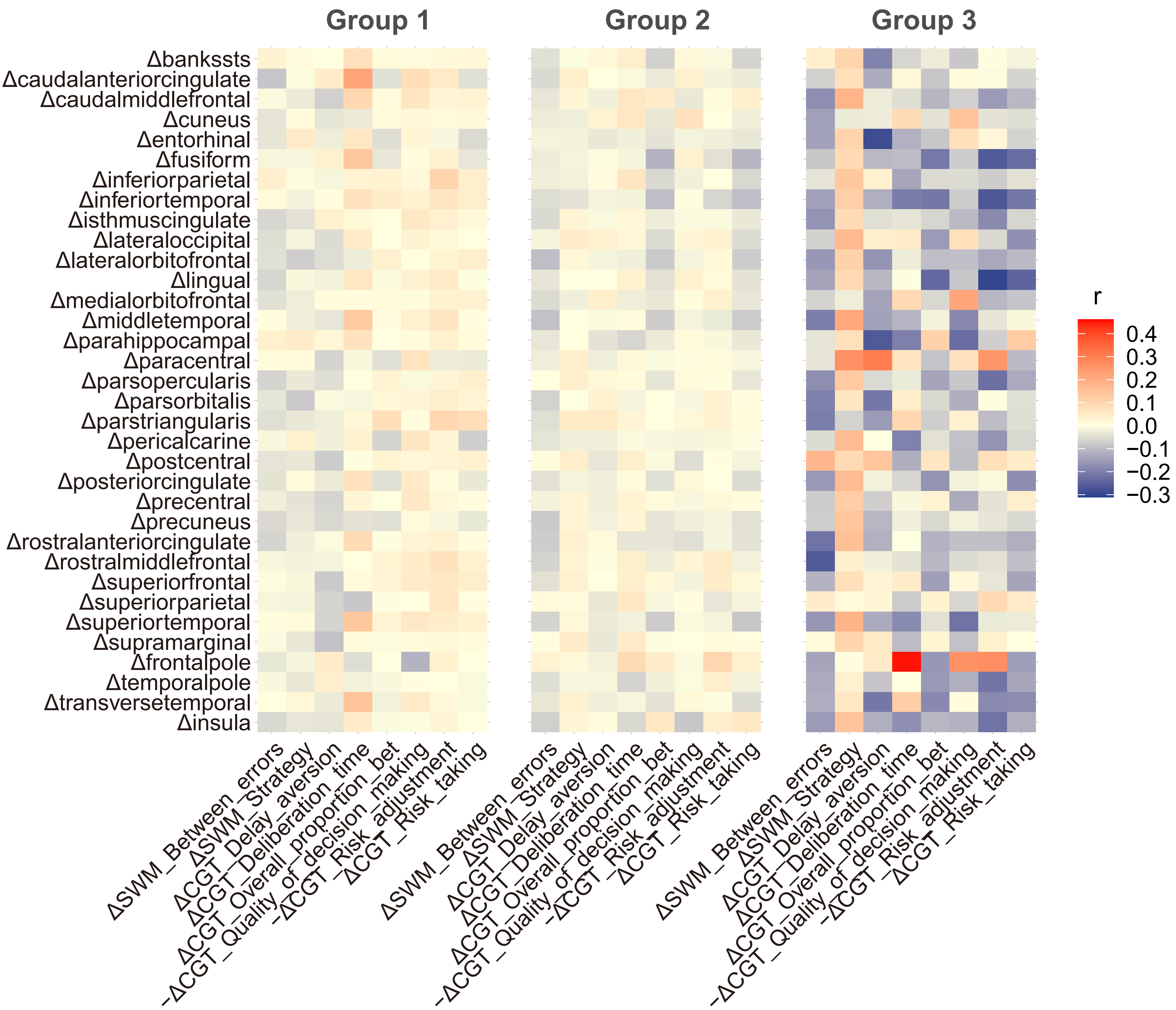


**Supplementary Fig. 8. Correlation between longitudinal trajectory of GMV in each cortical ROI and neurocognition in groups**. Negative correlations between the developmental trajectories of GMV in the top discriminating ROIs and neurocognitive performances for Group 2 (*n*=765) were observed while increasing GMV in the top discriminating ROIs showed positive correlation with improvements of neurocognitive functions for Group 3 (*n*=67).

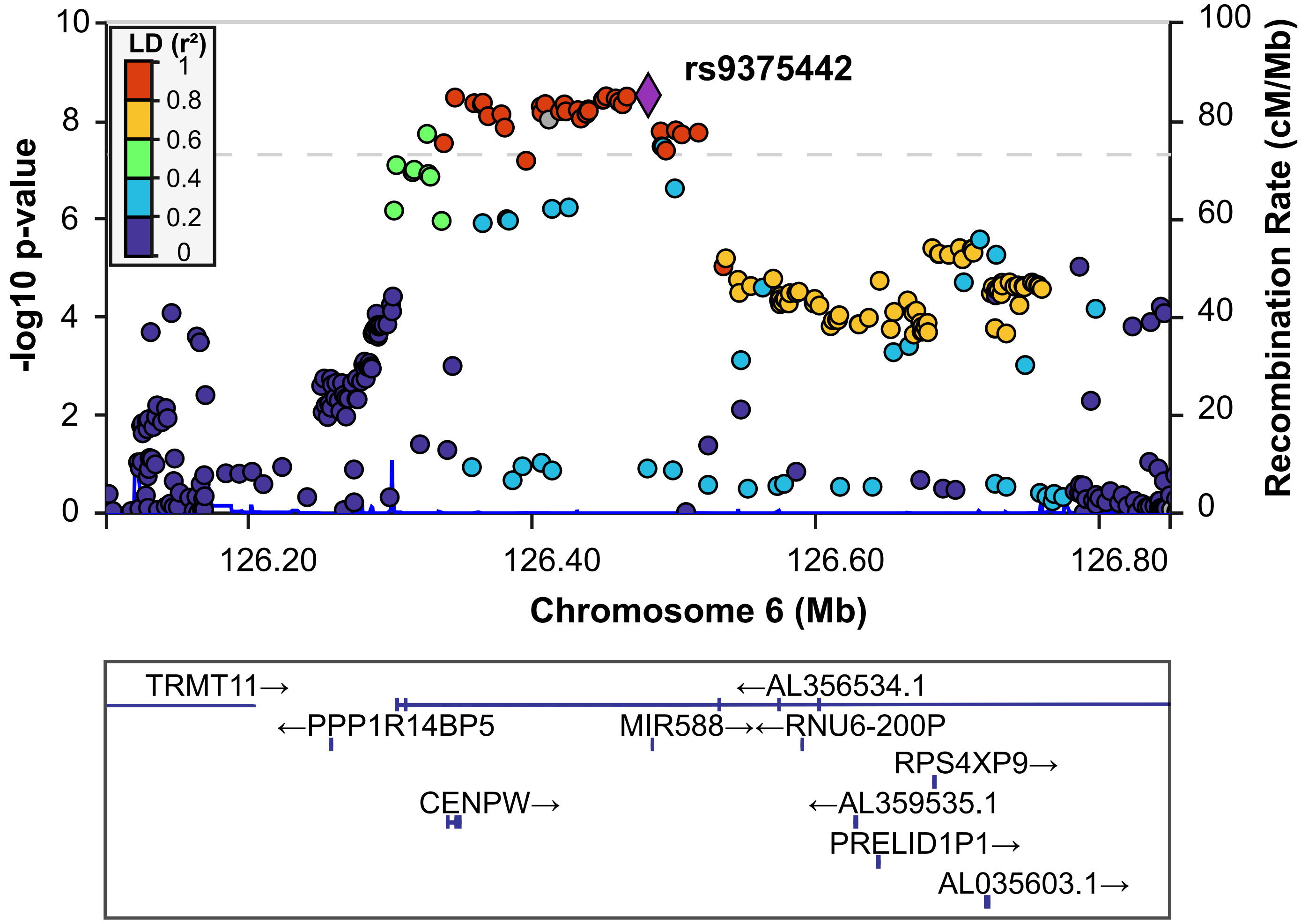


**Supplementary Fig. 9. Locus zoom of the top SNP rs9375442 on chromosome 6.**

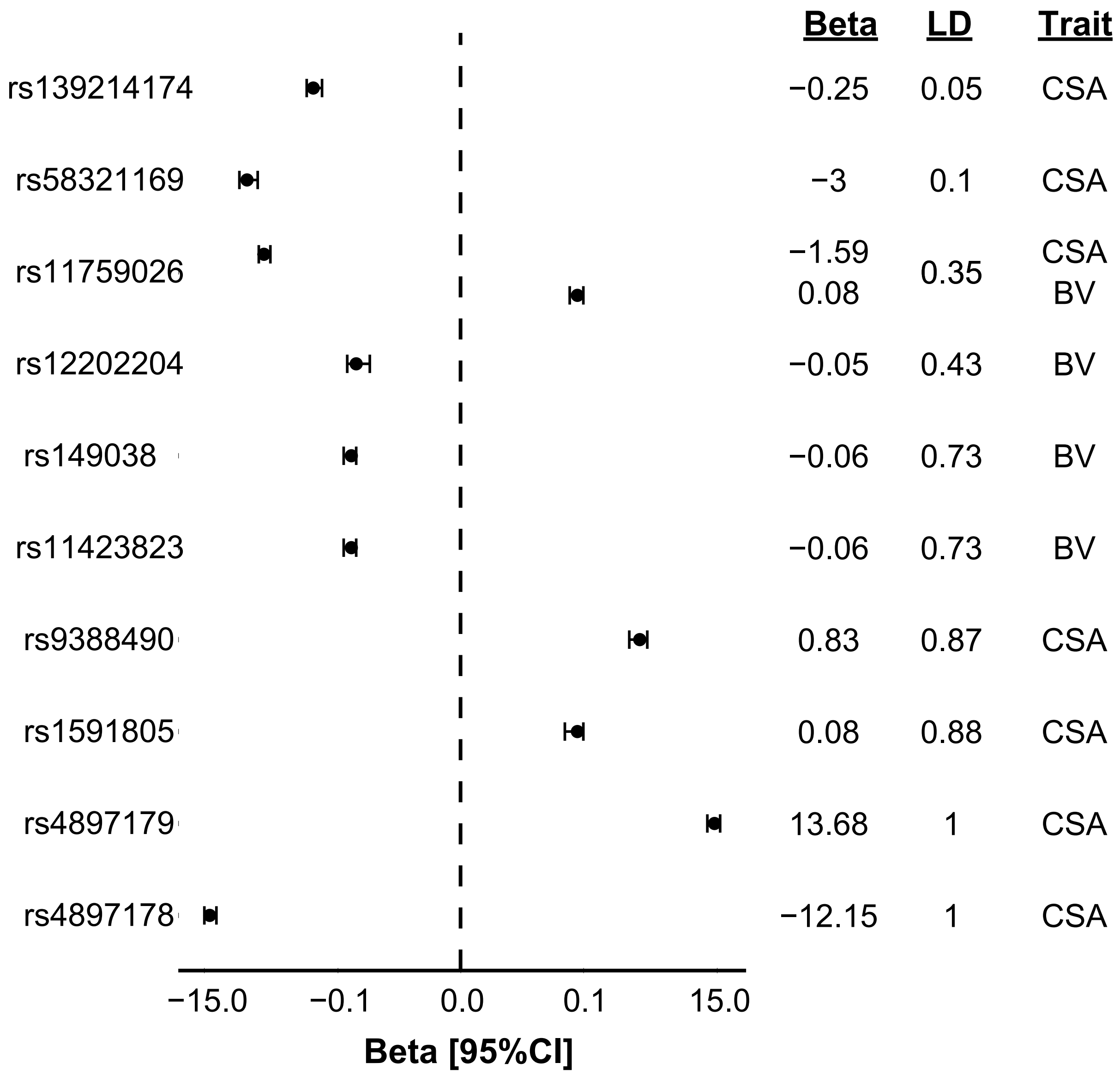


**eFigure 10. Associations between SNPs on CENPW and cortical surface area (CSA) and brain volume (BV) in existing studies.** Beta values and their 95% confidence intervals for each SNP were plotted in the forest plot, and LD were calculated for the corresponding SNP with the leading SNP from GWAS results in **Fig. 3b**.

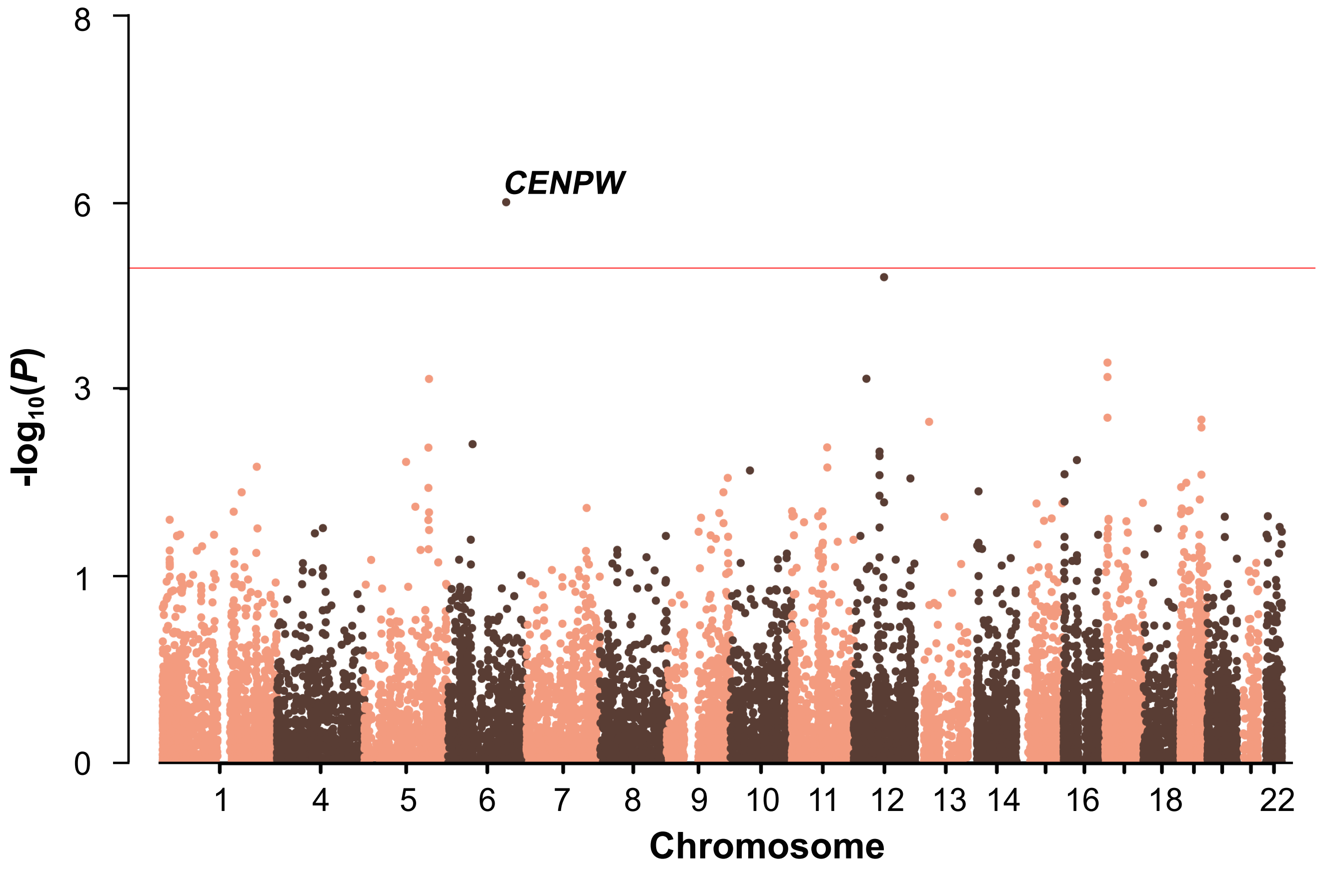


**Supplementary Fig. 11. Gene-based GWAS results for delayed neurodevelopment.** Gene-based association tests ($n$=7,662) identified one genome-wide significant genes (CENPW, $P$=9.86×$10^{-7}$) (significance level: $P$=5×$10^{-6}$) associated with delayed neurodevelopment in Group 3.

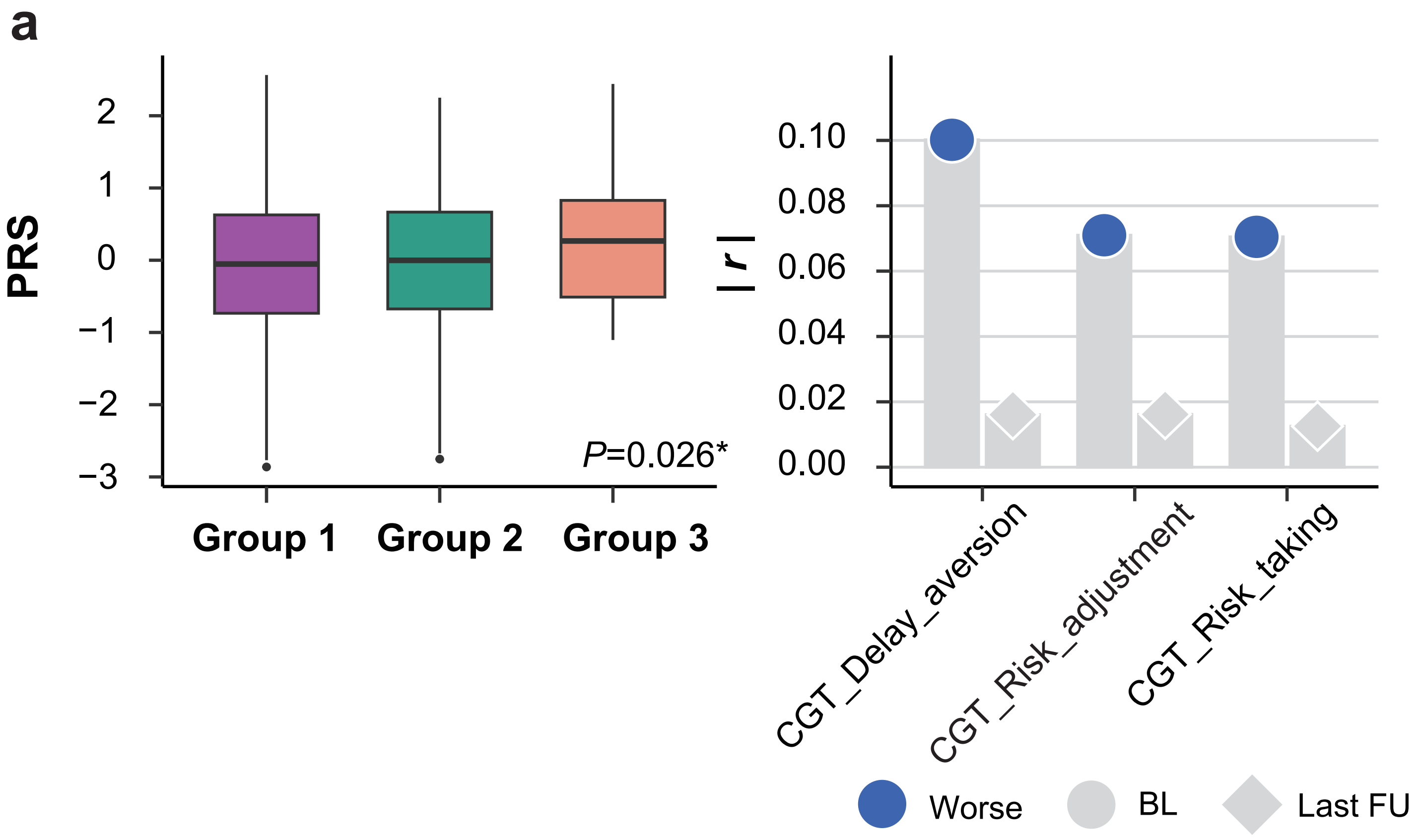


**Supplementary Fig. 12. Validation of GWAS results for delayed neurodevelopment in IMAGEN.** Box plot in **(a)** showed that PRS of delayed neurodevelopment was higher in Group 3 (*n*=60) compared to Group 1 and 2 (*n*=*1,338*) (two-sided t-test: *P*=0.012). **(b)** indicated that PRS of delayed neurodevelopment was negatively correlated with baseline neurocognitive performance, and became non-significant at the last follow-up. CGT delay aversion, BL ($r$=0.10, *$P_{adj}$=0.004), FU3 ($r$=-0.02, $P_{adj}$=0.682); CGT risk adjustment, BL ($r$=-0.07, *$P_{adj}$=0.032), FU3 ($r$=-0.02, $P_{adj}$=0.682); CGT risk taking BL ($r$=0.07, *$P_{adj}$=0.032), FU3 ($r$=0.01, $P_{adj}$=0.682).

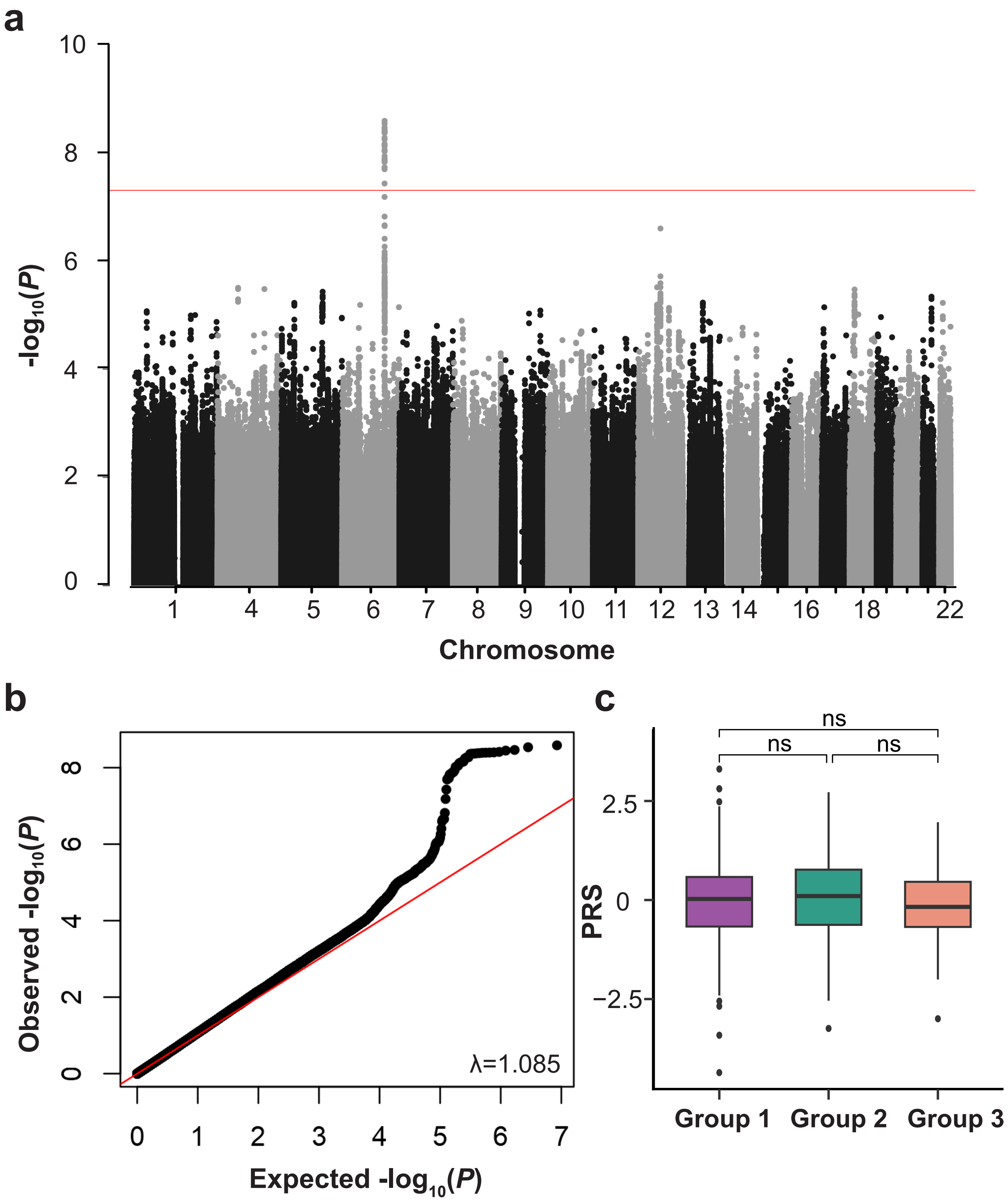


**Supplementary Fig. 13. GWAS results and validation for Group 2 vs Group 1.** (a) GWAS Manhattan plot for Group2-reweighted GMV in the ABCD population (*n*=7,662). Group2-reweighted GMV was calculated for each adolescent (**Methods**) and used as the proxy phenotype. (b) Q-Q plot of GWAS for Group 2 vs Group 1. (c) indicated that PRS of Group2-reweighted GWAS didn't differ among groups.

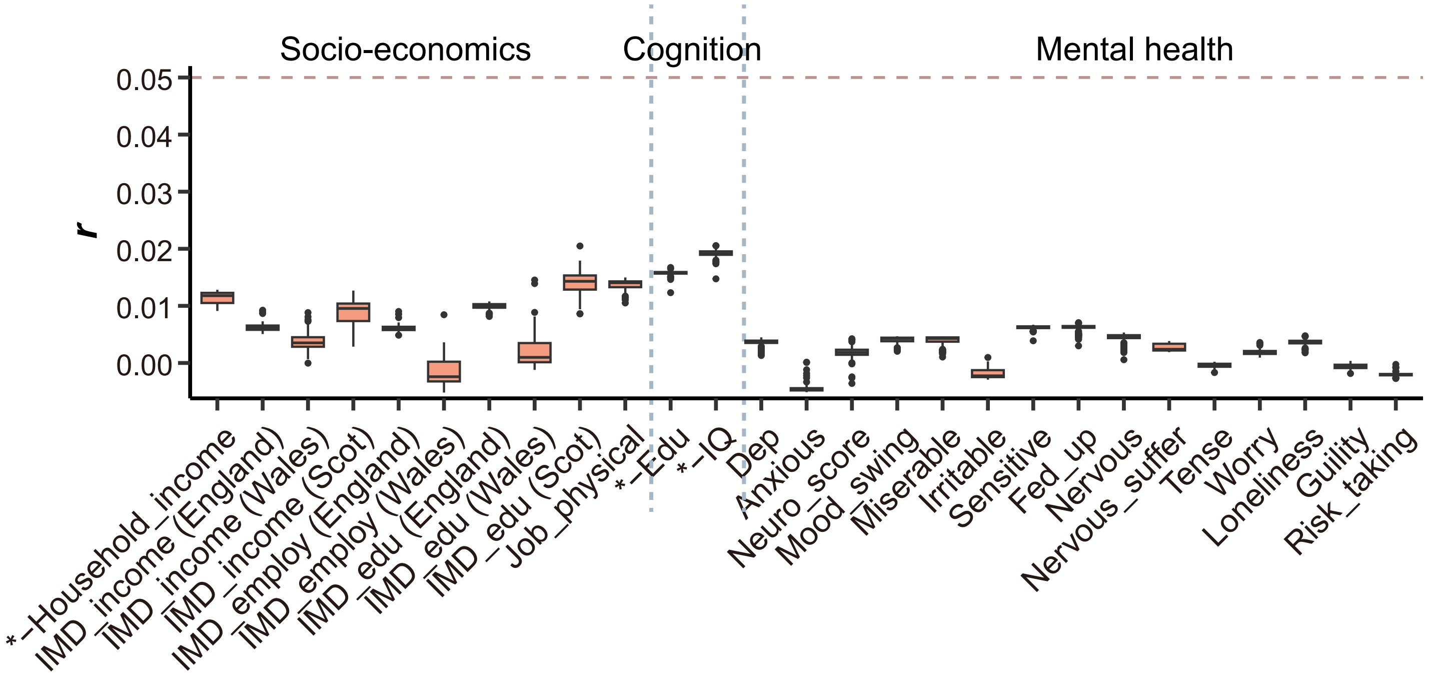


**Supplementary Fig. 14. Correlations between all PRS and socio-economics, cognitive and mental health and in UKB.** Non-inferiority test against 0.05 correlation coefficient showed that genetic variants had limited effect on the long-term cognitive, mental health and socio-economic outcomes (*n*=337,199).

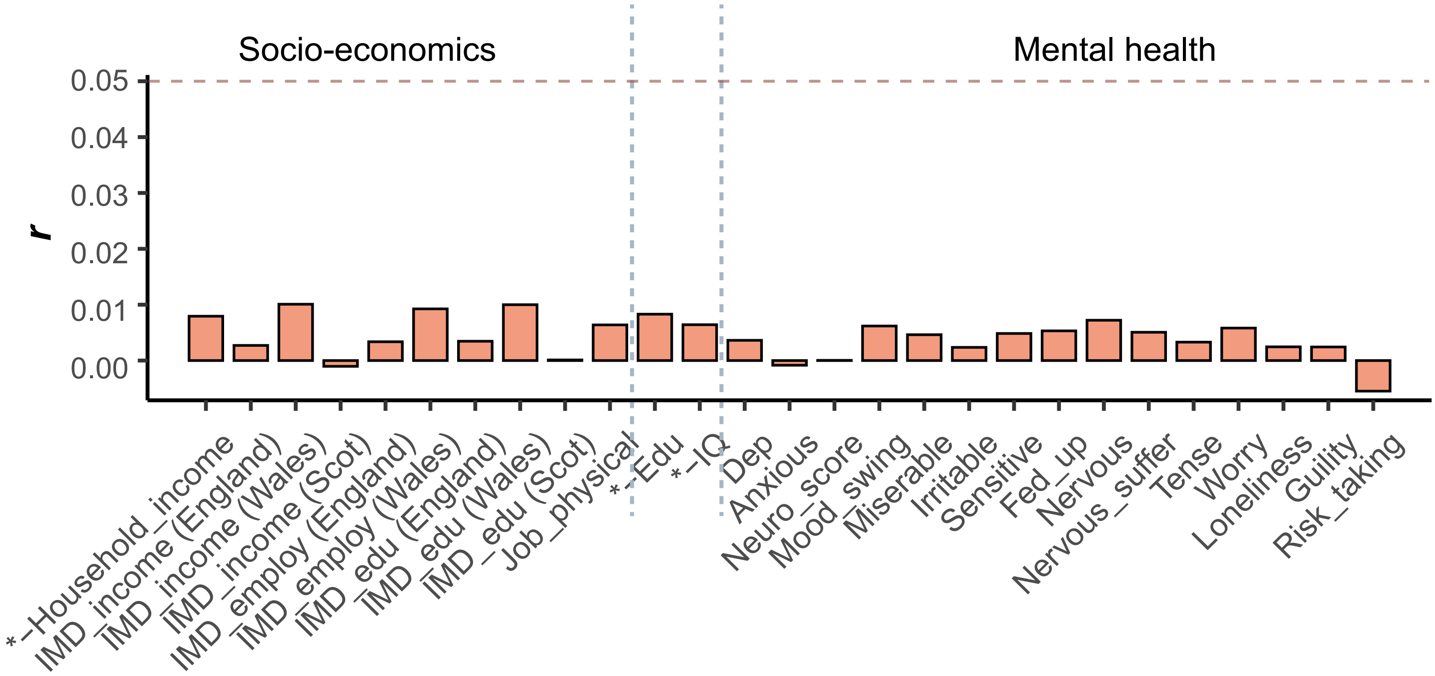


**Supplementary Fig. 15. Correlations between CENPW score and socio-economics, cognitive and mental health and in UKB.** Non-inferiority test against 0.05 correlation coefficient showed that genetic variants had limited effect on the long-term cognitive, mental health and socio-economic outcomes (*n*=337,199).

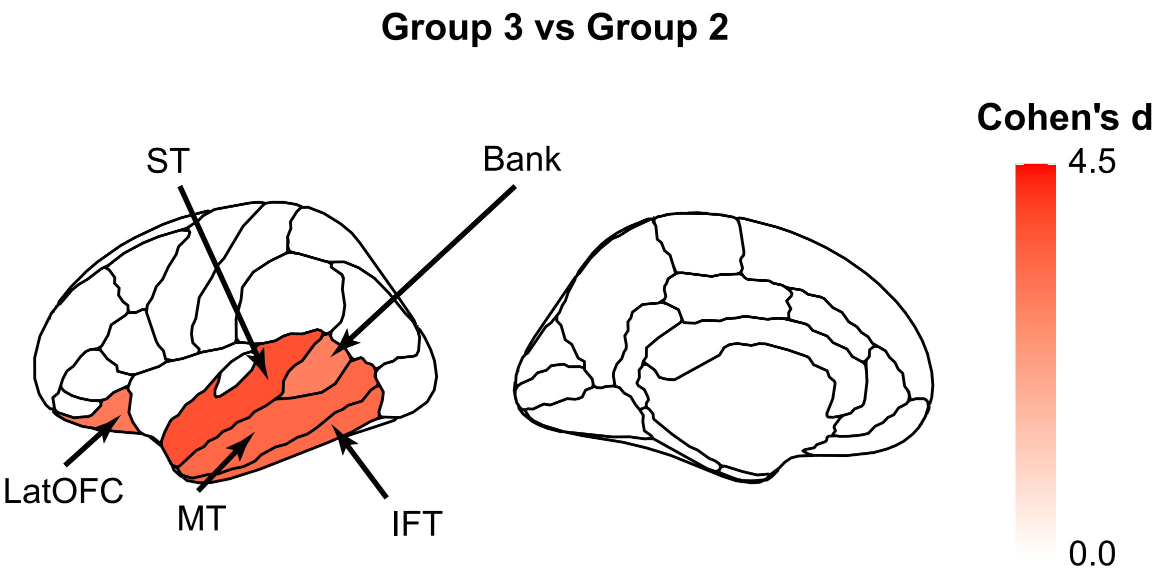


**Supplementary Fig. 16. The top 5 discriminating ROIs with largest t values comparing the GMV trajectories between Group 3 and Group 2**. Sex, imaging site, handedness and ICV were adjusted. IFT ($d$=3.33, $t$=16.65, ***$P_{adj}$<0.001), MT ($d$=3.37, $t$=16.28, ***$P_{adj}$<0.001), LatOFC ($d$=3.05, $t$=14.60, ***$P_{adj}$<0.001), ST ($d$=3.77, $t$=14.55, ***$P_{adj}$<0.001), Bank ($d$=*2.92*, $t$=14.50, ***$P_{adj}$<0.001). Bank, bankssts; IFT, inferior temporal; LatOFC, lateral orbitofrontal; MT, middle frontal; ST, superior temporal.

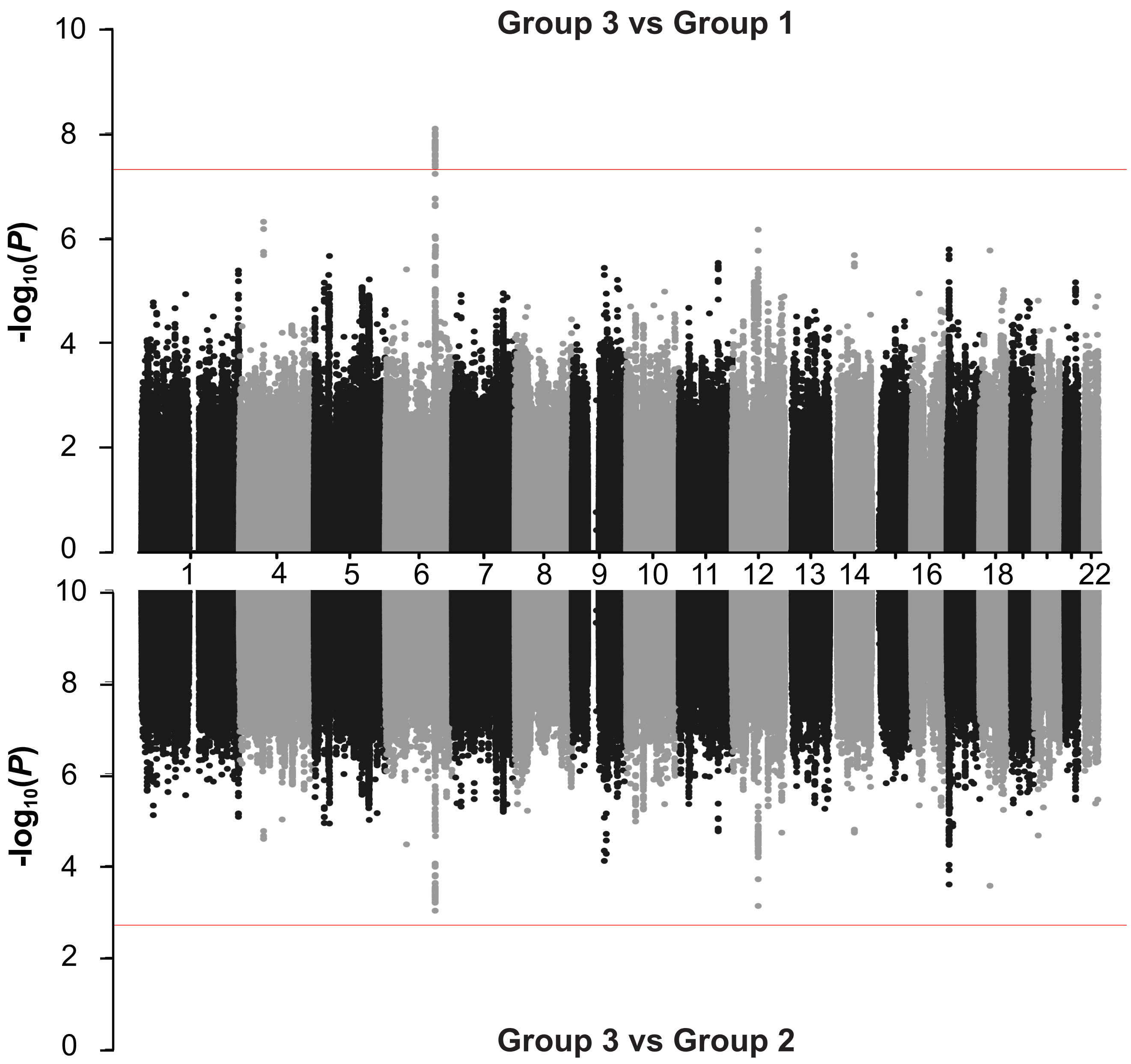


**Supplementary Fig. 17. Miami plot of GWAS for Group 3 vs Group 1 and Group 2.** GWAS Manhattan plots for Group3-reweighted (vs Group 1 on top and Group 2 on bottom) GMV in the ABCD population (*n*=7,662) were compared. Reweighted GMV was calculated for each adolescent (**Methods**) and used as the proxy phenotype.

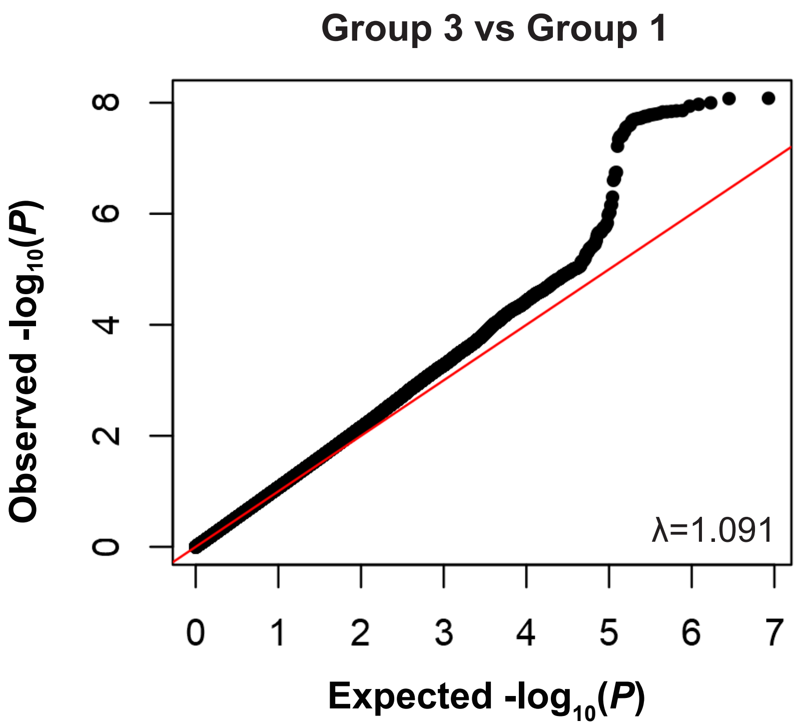


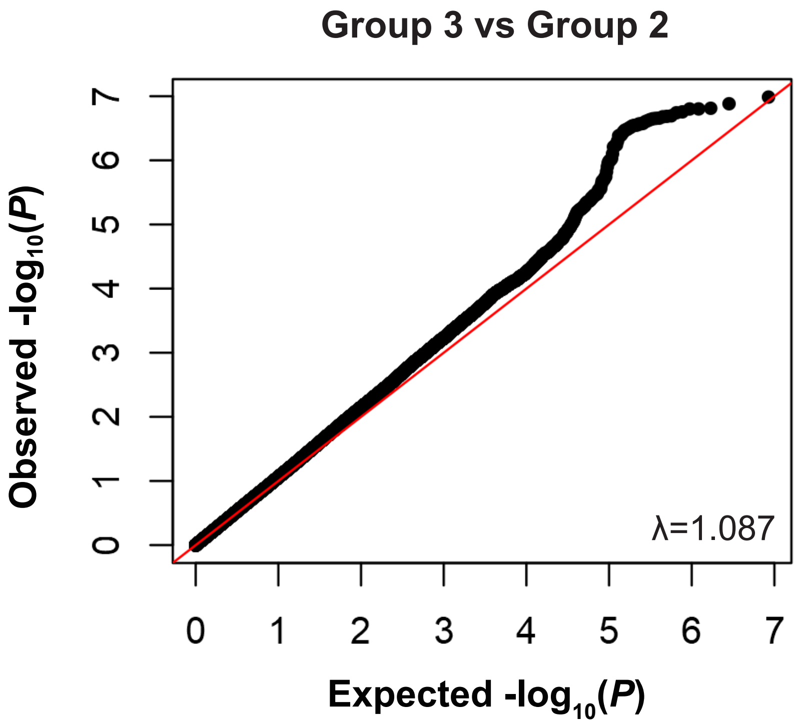


**Supplementary Fig. 18. Q-Q plots of GWAS for Group 3 vs Group 1 and Group 2.**

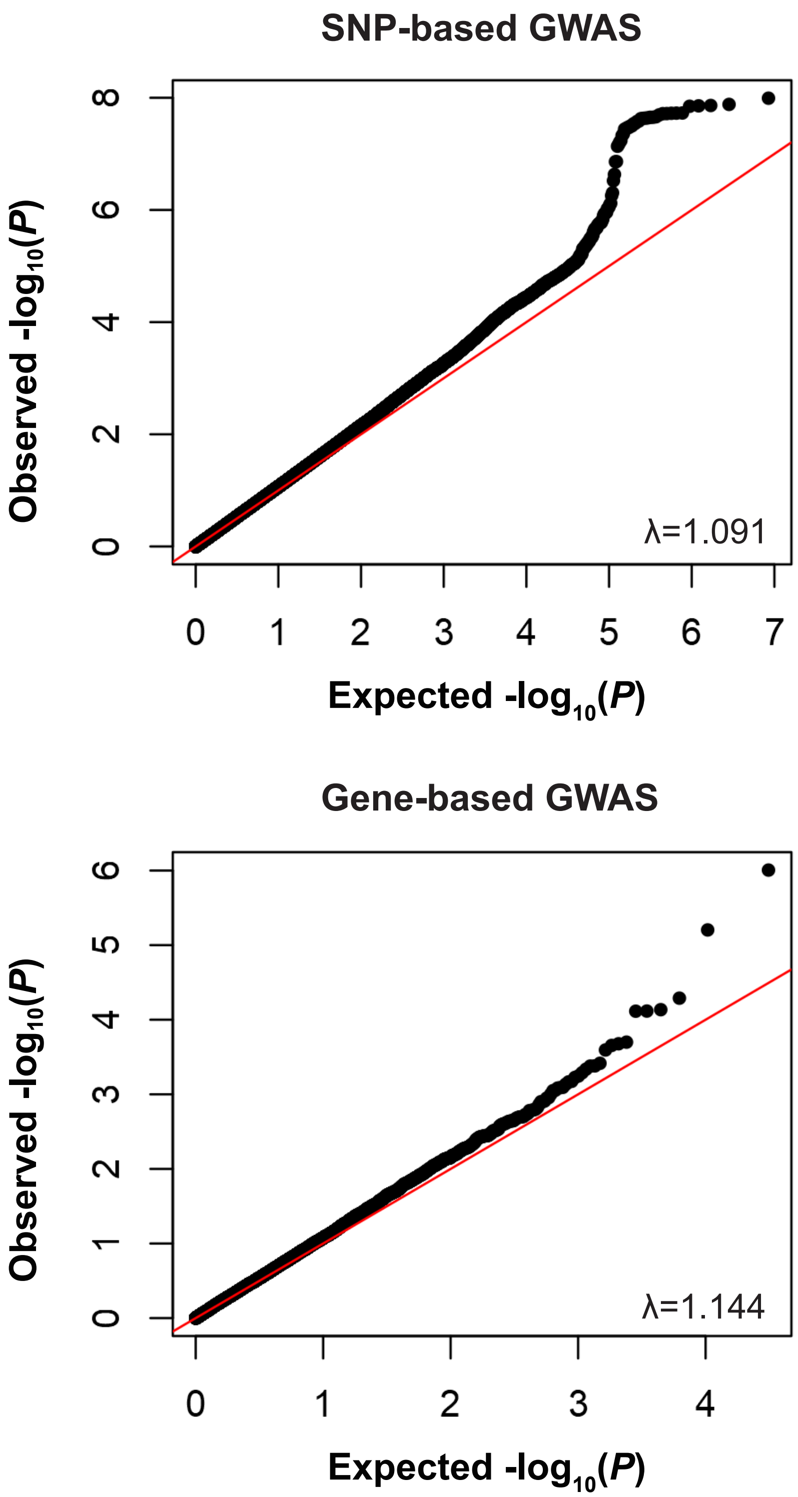


**Supplementary Fig. 19. Q-Q plots of GWAS for Group 3 vs Group 1/2.**
