## Supplementary Methods for "Structural neurodevelopment at the individual level - a life-course investigation using ABCD, IMAGEN and UK Biobank data"

### Description of population cohorts

<u>ABCD - Adolescent Brain and Cognitive Development</u>

The ABCD Study is a landmark, longitudinal study of brain development, examining approximately 11,875 youth from 21 sites across the United States from age 9 to 10 for approximately ten years into young adulthood[1]. Data from the 3.0 (for baseline data collected between 2016-2018, ages 9-11) and latest 4.0 (for follow-up data collected between 2017-2020, ages 11-13) annual curated data releases of the ABCD project (https://abcdstudy.org/about/) were included. In our analysis, the ABCD data serves two main purposes. Firstly, it is used to estimate the developmental curve of total gray matter volume (GMV) in the reference population and group-specific GMV developmental curve. Secondly, it is used to examine the genetic influence on distinct neurodevelopmental patterns during adolescence through genome-wide association study (GWAS). A total of 11,811 participants (11,760 were included at baseline) aged 8.92 to 13.83 years old with neuroimaging data available was included in the study, 7,776 (65.84%) of which had two scans. Among the participants, 6170 (52.24%) are males and 5641 (47.76%) are females.

*Structural neuroimaging data.*

The ABCD neuroimaging data were obtained using 3T scanners (Siemens Prisma, General Electric MR750 and Philips Achieva dStream) with 32-channel head coil and high resolution T1-weighted structural magnetic resonance imaging (MRI). Both the methods and evaluations of these MRI images have been harmonized and optimized across all ABCD research sites[2,3]. The pre-processing processes were completed by the ABCD research teams according to the ABCD standard pipeline and protocol, with details described in the image processing paper[3]. The quality control (QC) procedure of the processed neuroimages was checked by the ABCD team both automatically and manually. Then regional morphometric structure evaluations were obtained using FreeSurfer 6.0 including cortical volumes from the FreeSurfer Desikan-Killiany (h.aparc) atlas, and subcortical volumes from the ASEG atlas. According to the FreeSurfer reconstruction QC measures (freesqc01), a total of 1,9576 scans including 11,811 participants passed the QC were included in the structural analyses. As there is more than one scanner in several sites, we controlled for scanner (*mri_info_deviceserialnumber* variable in *abcd_mri01* file) rather than site in the analysis. A total of 31 scanners was used in ABCD. Because most of the sample size of each scanner was large enough, regression method is able to substantially mitigate scanner effects.

*Genetic data*

The ABCD imputed genotype data were obtained from the public release 3.0. Imputation was performed using the Michigan Imputation Server with hrc.r1.1.2016 reference panel[4] and Eagle v2.3 phasing. We performed stringent QC standards by PLINK 1.90. Individuals with >10% missing rate and single-nucleotide polymorphisms (SNPs) with call rates < 95%, minor allele frequency < 0.1%, deviation from the Hardy-Weinberg equilibrium with P < 1E-10 were excluded from the analysis, yielding 11,1014 participants and 244,227 SNPs. To ensure the homogeneity of the ABCD and IMAGEN population, we selected only ABCD subjects self-reporting ancestral origins as white, with 2,387 participants excluded. Considering that ABCD is oversampled for siblings and twins, and thereby has a nested structure, we randomly selected one participant within a family (the kinship relationship between participants was decided by genetically inferred zygosity status in *acspsw03* file). Finally, a total of 7,662 participants was included in the genetic analysis.

<u>IMAGEN</u>

The IMAGEN study is a significant multicenter genetic-neuroimaging study aimed to investigate the genetic and neurobiological basis of individual variability in neurocognition, and determining their predictive value for the development of frequent psychiatric disorders[5]. The study recruited approximately 2,000 healthy Caucasian adolescents at age 14 (BL) from middle-class school across Europe in 8 sites (Berlin, Dresden, Dublin, Hamburg, London, Mannheim, Nottingham and Paris). Out of the recruited participants, 2,138 participants had neuroimaging data available. At each evaluation, participants had a structural MRI scan and a comprehensive assessment of their individual, social and family characteristics. Adolescents were followed up at the age of 16 years (FU1), 19 years (FU2) and 23 years old (FU3). However, neuroimaging data were only available at BL, FU2 and FU2. A total of 1,543 participants was included in the analysis. Among them, 747 (48.41%) are males and 796 (51.59%) are females.

*Structural neuroimaging data*

High-resolution T1-weighted images were obtained using 3T MRI systems based on the ADNI protocol (http://www.loni.ucla.edu/ADNI/Cores/index.shtml), from 4 different manufacturers (Siemens: 4 sites, Philips: 2 sites, General Electric: 1 site, and Bruker: 1 site). The scanning variables were specially chosen to be compatible with all scanners. The MR protocols and QC procedures of the IMAGEN study are described in Schumann et al.[6]. In brief, preprocessing is performed centrally using an automated pipeline that processes the continuously incoming data, and accounts for inter-site variability. In addition to the standard IMAGEN procedures, FreeSurfer (v6.0) Desikan-Killiany and ASEG atlas were used to extract regional brain morphology. 2,138 participants with 4,681 scans passed the QC. 21 individuals with GMV beyond 4 interquartile ranges (IQRs) in any left/right hemisphere regions were considered to be outliers and were excluded from the following analyses. To ensure a more accurate estimation of individual trajectories from linear mixed effect model, only 1,543 participants with at least two MRI scans were included in the study. Among them, 974 adolescents having a total of 3 scan and 569 adolescents having a total of 2 scans. As there is only one scanner manufacturer in every site, site was used as a covariate to control for potential scanner effect.

*Genetic data and epigenetic data*

Details of the genotyping and quality control are available in Desrivières et al.[7]. A same QC processing as the one in ABCD was performed using PLINK 1.90, where SNPs with call rates < 95%, minor allele frequency < 0.1%, deviation from the Hardy-Weinberg equilibrium with P < 1E-10 were excluded from the analysis. Then an imputation was conducted on the quality-controlled genetic data using the TOPMed imputation server (https://imputation.biodatacatalyst.nhlbi.nih.gov) with the HapMap3 reference panel[8]. After imputation, 5,966,316 SNPs were available for 1,982 IMAGEN sample. Among them, 1,398 were included in the analysis (644 for group 1, 694 for group 2 and 60 for group 3).

Epityping was conducted on DNA extracted from peripheral blood cells using the Infinium HumanMethylation450K BeadChip (Illumina) running on an Illumina HiScan System using the manufacturer's standard protocol. Pyrosequencing was carried out for technical validation and independent replication with a PSQ96 genetic sequencer using PyroMark Gold Q96 reagents (Qiagen, Valencia, Calif.) in accordance with the manufacturer's recommendations. DNAm beta values were normalized using Illumina GenomeStudio software. Quality control was performed by excluding CpGs with detection P < 0.01 and including samples had >98% sites with detection P < 0.01, yielding 372,582 CpGs and 1,329 samples. Among them, 446 were identified as group 1, 463 were identified as group 2 and 36 were identified as group3.

HCP - Human Connectome Project

The HCP consortium shared several large-scale cross-sectional neuroimaging datasets, which can be accessed through the HCP website (https://www.humanconnectome.org), including HCP Development (HCP-D) and HCP Young Adult (HCP-YA). The details on the inclusion and exclusion criteria of HCP YA were provided in the previous study[9]. HCP-YA sampled 300-400 healthy young adult sibships of average size 3-4, with most of these sibships including a MZ or DZ twin pair. A total of 1,113 participants from 457 unique families (including 170 dizygotic twins, 286 monozygotic twins, 576 non-twin siblings, and 25 non-sibling familial relations) aged 22-37 years old was included in the analysis with neuroimaging data available, comprising of 507 (45.55%) and 606 (54.45%) females. HCP-D is an extension of the HCP-YA study, specifically aimed at extending the coverage of HCP to a fuller lifespan[10]. It follows a comparable acquisition and recruitment protocol to HCP-Y but focuses on younger participants. A total of 652 participants aged 5.58-21.92 years old was included in the analysis with neuroimaging data available, comprising of 301(46.17%) males and 351 (53.83%) females.

*Structural neuroimaging data*

HCP-YA imaging data were acquired on a Siemens Skyra 3T scanner employing a 32-channel head coil with a customized SC72 gradient insert. HCP-D imaging data is conducted on a 3T Siemens Prisma scanners (Siemens, Erlangen, Germany). Participants 8-21 years old are scanned using the Siemens 32-channel Prisma head coil; a pediatric 32-channel head coil developed by Ceresensa (www.ceresensa.com) is used for 5-7 years old participants (see Harms et al. (under review)). Besides, contrast to HCP-YA, a slightly larger T1w voxel size (0.8mm) to allows some additional SNR margins, use of volumetric navigators for prospective motion correction, only one acquisition per modality to reduce scanning time, multi-echo acquisition for T1 (TE=1.8, 3.6, 5.4 and 7.2ms), slower TR (800ms) to allows maintenance of full Fourier k-space acquisition necessitated by the increased number of echoes were used for HCP-D participants. The preprocessing in HCP-D follows a similar procedure as in HCP-Y. Detailed protocols are available at https://www.humanconnectome.org. Minimally processed data was obtained directly from HCP and then be used to extract regional volumes using Freesurfer 6.0 Desikan-Killiany and ASEG atlas.

PNC - Philadelphia Neuroimaging Cohort

PNC is a large-scale study of child development that incorporates rich multi-modal neuroimaging, genetics, and detailed clinical and cognitive phenotyping. The PNC data includes information from 9,498 children recruited from the Children's Hospital of Philadelphia care network. A sub-sample including approximately 1,000 healthy participants received multi-modal neuroimaging performed on a separate study visit at Penn[11]. A total of 1,587 participants aged 8.08-23.08 years old was included in the analysis with neuroimaging data available, comprising of 756 (47.64%) males and 831 (52.36%) females.

*Structural neuroimaging data*

All MRI scans were acquired at a single site, on a single 3T Siemens TIM Trio scanner with 32-channel head coil, in a short period of time that did not span any software or hardware upgrades as described previously[12]. In brief, receive coil shading was reduced by selecting the Siemens prescan normalize option, which corrects for B1 inhomogeneity based on a body coil reference scan. Image quality assessment was performed using visual inspection, which primarily focused on identifying excessive subject motion. Quality-controlled processed data was obtained directly from PNC and then be used to extract regional volumes using Freesurfer 6.0 Desikan-Killiany and ASEG atlas.

UKB - UK Biobank

UKB study provides a large, comprehensive and ongoing dataset that includes both extensive phenotypic information as well as neuroimaging and genetics with over 500,000 participants aged 40-69 years across UK when recruited in 2006-2010[13]. In this study, we only used the baseline data to assess the genetic-predicted risk of delayed neurodevelopment on long-term outcomes. Socioeconomic, cognition and mental health outcomes, and structural brain morphology at mid-to-late adulthood were of interest. A total of 502,409 participants aged 37-73 years old was included in the analysis, comprising of 229084 (45.60%) males and 273325 (54.40%) females.

*Structural neuroimaging data*

Structural minimally processed T1-data was collected by the UK Biobank study with identical hardware and software in Manchester, Newcastle, and Reading, collected with a standard Siemens Skyra 3T scanner with a 32-channel head coil. Brain volumetric phenotypes were pre-processed by an imaging-pipeline developed and executed on behalf of UK Biobank[14]. Details of the imaging protocol can be found in an open-source document (https://biobank.ndph.ox.ac.uk/showcase/showcase/docs/brain_mri.pdf). Volumetric measures ($mm^3$) have been generated in each participant's native space. We used imaging-derived phenotypes of cortical and subcortical grey-matter volumes in regions of interest (UK Biobank category 192 & 190). A total of 43,103 participants balanced between sex was available with sMRI data across 22 sites.

*Genetic data*

Genotype data were available for all the initial participants in the UK Biobank cohort. Detailed genotyping and quality control procedures for the UK Biobank are available in a previous publication[15]. Additional QC procedures were conducted the same as described in ABCD and IMAGEN. We excluded SNPs with call rates < 95%, minor allele frequency < 0.1% or deviation from the Hardy-Weinberg equilibrium with $P < 1E-10$ and selected individuals that were estimated to have recent British ancestry and have no more than ten putative third-degree relatives in the kinship table, yielding 616,339 SNPs and 337,199 participants.

Table SM1. A sketch of data utilization in analysis.

| | ABCD | IMAGEN | UKB | HCP | PNC |
|---|---|---|---|---|---|
| Group identification and characterization | | √ | | | |
| Estimation of group-specific GMV developmental curve | √ | √ | | √ (HCP-D) | √ |
| Estimation of peak total GMV in IMAGEN | √ | √ | | √ (HCP-D / YA) | √ |
| GWAS and GWAS validation | √ | | √ | | |
| EWAS and EWAS validation | | | √ | | |
| Long-term impacts of delayed neurodevelopment | | √ | | | |

### Assessment instruments in the ABCD, IMAGEN and UKB.

Table SM2. Neurocognition assessments in ABCD

| Instrument | Variable | Description |
|---|---|---|
| Game of Dice Task[16] | gdt_scr_values_safe | Counts how many times participants selected a safe bet (bets on 3 or 4 dice faces) |

| | | |
|---|---|---|
| | gdt_scr_values_risky | Counts how many times participants selected a risky bet (bets on 1 or 2 dice faces) |
| Delay Discounting Task[17] | ddis_scr_expr_mnrt_immcho / ddis_scr_expr_mnrt_allcho | mean latency of 'immediate' choices adjusted by mean latency of all choices |
| | ddis_scr_expr_mnrt_delaycho / ddis_scr_expr_mnrt_allcho | mean latency of 'delayed' choices adjusted by mean latency of all choices |
| NIH Tool Box[18] | | |
| Picture Vocabulary Test | nihtbx_picvocab_uncorrected | a measure of general vocabulary knowledge |
| Flanker Inhibitory Control and Attention Test | nihtbx_flanker_uncorrected | a measure of executive function, specifically tapping inhibitory control and attention. |
| List Sorting Working Memory Test | nihtbx_list_uncorrected | a measure of episodic memory |
| Dimensional Change Card Sort Test | nihtbx_cardsort_uncorrected | a measure of executive function, specifically tapping cognitive flexibility |
| Pattern Comparison Processing Speed Test | nihtbx_pattern_uncorrected | a measure of speed of processing |
| Picture Sequence Memory Test | nihtbx_picture_uncorrected | a measure of episodic memory |
| Oral Reading Recognition Test | nihtbx_reading_uncorrected | a measure of reading decoding skill |
| Fluid Cognition | nihtbx_fluidcomp_uncorrected | A composite of Flanker, Dimensional Change Card Sort, Picture Sequence Memory, List Sorting and Pattern Compariso, which plays an important role in adapting to novel situations in everyday life |
| Crystallized Cognition | nihtbx_cryst_uncorrected | A composite of Picture Vocabulary and Reading Tests, which represents an accumulated store of |

| | | |
|---|---|---|
| | | verbal knowledge and skills |
| Total Cognition | nihtbx_totalcomp_uncorrected | A combination of fluid and crystallized composites |

Table SM3. Environmental, neurocognition, behavioral, personal trait and mental health assessments in IMAGEN

| Instruments | Variables | Descriptions |
|---|---|---|
| Environmental factors | | |
| Life Events Questionnaire[19] (LEQ) | Family/Parents | Mean life-time frequency of events comprising parental divorce, parental discord, parental remarriage, parental alcohol abuse, family financial difficulties |
| | Accident / Illness | Mean life-time frequency of events comprising accident/illness, given medication by a physician, death in family, serious accident or illness |
| | Sexuality | Mean life-time frequency of events comprising falling in love, starting or ending a relationship, having first sexual experience, having a gay experience, pregnancy |
| | Autonomy | Mean life-time frequency of events related to independence including starting college, a hobby or new friends |
| | Deviance | Mean life-time frequency of events comprising getting in trouble, at school or the law, stealing |
| | Relocation | Mean life-time frequency of events related to change of school or residence |
| | Distress | Mean life-time frequency of events comprising face breaking out with pimples, starting to see a therapist, thinking about suicide, running away from home, getting poor grades at school, and gaining a lot of weight |
| | Overall valence | Mean life-time frequency of all measured events |

| | | |
|---|---|---|
| Childhood Trauma Questionnaire[20] (CTQ) | Emotional abuse | Verbal assaults on a child's sense of worth or well-being or any humiliating or demeaning behavior directed toward a child by an adult or older person (CTQ_3+ CTQ_8+ CTQ_14+ CTQ_18+ CTQ_25) |
| | Physical abuse | Sexual contact or conduct between a child younger than 18 years of age and an adult or older person (CTQ_9+CTQ_11+CTQ_12+CTQ_15+CTQ_17) |
| | Sexual abuse | Bodily assaults on a child by an adult or older person that posed a risk of or resulted in injury. (CTQ_20+CTQ_21+CTQ_23+CTQ_24+CTQ_27) |
| | Emotional neglect | The failure of caretakers to meet children's basic emotional and psychological needs, including love, belonging, nurturance, and support (*CTQ_5+*CTQ_7+*CTQ_13+*CTQ_19+*CTQ_28) |
| | Physical neglect | The failure of caretakers to provide for a child's basic physical needs, including food, shelter, clothing, safety, and health care (CTQ_1+*CTQ_2+CTQ_4+CTQ_6+CTQ_26) |
| Development Well-being Assessment Interview (DAWBA)[21] - Family Stress Scale | Socioeconomics | Family stressors in socioeconomic/housing |
| | Work | Family stressors in work/pressure |
| | Health | Family stressors in illness |
| | Addiction | Family stressors in relationships/addition |
| DAWBA[21] - Family Life Questionnaire | Affirmation | Behaviors that the parent puts in place to support or help children in various situations or to show them approval and affection and refers to parent-child relationship (i.e. 'gets love and affection', 'praised and rewarded', 'gets help and support when stressed' and 'liked and respected') |
| | Discipline | Behaviors by parents in response to and intended to |

| | | |
|---|---|---|
| | | correct misbehavior by the children and it refers to punishment (i.e. 'physical punishment' and 'non-physical punishment') |
| | Rules | The ability to create coherent and shared family rules and to enforce them and it measures structure and organization within the family (i.e. 'clear rules' and 'consistently applied rules') |
| | Special allowance | Overprotection behaviors as opposed to lack of supervision (i.e. 'very protected' and 'spends time alone') |
| Neurocognition | | |
| Cambridge Neuropsychological Test Automated Battery[22] (CANTAB) | | |
| Pattern recognition memory (PRM) - visual pattern recognition memory | Percent correct | Higher the percentage of correct trails, better the visual pattern recognition memory |
| Affective Go/No Go Task (AGN) - decision making | Total omissions for positive category | The AGN test assesses information processing biases for positive and negative stimuli. The subject is given a target category, and is asked to press the press pad when they see a word matching this category |
| | Total omissions for negative category | |
| Spatial Working Memory (SWM) - executive function | Between error | The number of times touching boxes that have been found to be empty and revisiting boxes which have already been found to contain a token |
| | Strategy | The number of times a subject begins a new search with a different box for 6- and 8-box problems only. A high score represents poor use of this strategy and a low score |

| | | |
|---|---|---|
| | | equates to effective use |
| Cambridge Gambling Test (CGT) - decision making | Delay aversion | Tendency to bet larger amounts when possible bet amounts are presented in descending order, calculated by subtracting risk taking measure from ascending trials from risk taking measure of descending trials |
| | Deliberation time | Mean latency from presentation of colored boxes to subject's choice of which color to bet on |
| | Overall proportion bet | Average proportion of the current point total (using nominal percentage) that subject is willing to risk on each gamble test trial |
| | Quality of decision making | Proportion of test trials on which the subject bets on the more likely outcome of the two choices |
| | Risk adjustment | Tendency to bet higher proportion of points when the large majority of boxes are the color chosen |
| | Risk taking | Mean proportion of the current points total that subject is willing to risk on trial for which they have chosen the more likely outcome |
| Rapid Visual Information Processing (RVP) - attention | A | A signal detection theory measure of target sensitivity |
| Intra-Extra Dimensional Set Shift (IED) - attentional flexibility | Total trials | The number of trials completed on all attempted stages |
| | Total trials adjusted | Attempts to compensate for the fact that subjects failing at any stage of the test have had less opportunity to complete trials. The adjustment adds 50 for each stage not attempted due to failure at an earlier stage |
| | PreED errors | The number of errors made prior to the extra-dimensional shift of the task. Errors are defined as instances when the subject fails to select the stimulus that is compatible with the current rule |

| | | |
|---|---|---|
| | ED errors | The number of errors made in the extradimensional stage of the task. Errors committed at the reversal stage following the EDS stage are not included |
| | Total errors | The number of errors in all 9 stages |
| | Total errors adjusted | The adjustment adds 25 for each stage not attempted due to failure at an earlier stage |
| Monetary-Choice Questionnaire[23] (KIRBY) - impulsivity | Estimated K | A measure of delay discounting. Larger k values indicating greater delay discounting of value for the delayed options, indicating a higher level of impulsivity |
| | Estimated K for small LDRs | K value for small long delay reward |
| | Esitmated K for Medium LDRs | K value for small long delay reward |
| | Estimated K for Large LDRs | K value for small long delay reward |
| Stop Signal Task[24] (SST) | Go reaction time | Mean response time for 'Go' trials |
| | Go accuracy | Rate of correct "Go" trials |
| | Stop accuracy | Rate of correct "Stop" trials |
| | Stop signal reaction time | A measure indicative of the inhibitory control process with a lower value indicative of better such control |
| Behavioral risk factors | | |
| DAWBA - Strengths and Difficulties Questionnaire[25] (SDQ) | Total difficulties score | Summing scores from all the scales except the prosocial scale |
| | Emotion problems score | i.e. "Many worries", "Nervous or clingy in new situations" |
| | Conduct problems score | i.e. "Often has temper tantrums or hot tempers", "Often lies or cheats" |
| | Hyperactivity score | i.e. "Restless, overactive", "Easily distracted, concentration wanders" |

| | | |
|---|---|---|
| | Peer problems score | i.e. "Rather solitary, tends to play alone", "Picked on or bullied by other children" |
| | Prosocial score | i.e. "Considerate of other people's feelings", "Often volunteers to help others" |
| | Impact score | The sum of distress and impairment scores (i.e. interfere with home life/friendships/classroom learning/leisure activities by self-report) |
| European School Survey Project on Alcohol and Drugs[26] (ESPAD); Fagerstrom test for nicotine dependence[27] (FTND) | Lifetime smoking | "On how many occasions during your lifetime have you smoked cigarettes?" |
| | smoking last month | "How frequently have you smoked cigarettes during the last 30 days?" |
| | Whole life drink | "On how many occasions in your whole lifetime have you had any alcoholic beverage to drink?" |
| | Drink last year | "On how many occasions over the last 12 months have you had any alcoholic beverage to drink?" |
| | Drink last month | "On how many occasions over the last 30 days have you had any alcoholic beverage to drink?" |
| Personal trait | | |
| NEO Five-Factor Inventory[28] | Neuroticism | Tend to accept negative emotions all the time, ignoring all the positive factors in life |
| | Extraversion | Like to engage with new people in social surroundings and are found to strengthen relationships |
| | Openness | Be open to new experiences and are often named under challenging personality |
| | Agreeableness | Be altruistic, sympathetic, and cooperate with everyone |
| | Conscientiousness | Be extensive with their duties and responsibilities |
| Temperament and Character Inventory[29] (TCI) - novelty seeking | Exploratory excitability | vs. stoic rigidity (i.e. "I prefer to start conversations, rather than waiting for others to talk to me") |
| | Impulsiveness | vs. reflection (i.e. "I often follow my instincts, hunches, |

| | | |
|---|---|---|
| scale | | or intuition without thinking through all the details") |
| | Extravagance | vs. reserve (i.e. "I prefer spending money rather than saving it") |
| | Disorderliness | vs. regimentation (i.e. "I lose my temper more quickly than most people") |
| | Total Novelty Seeking score | A sum of scores from the above four subscales |
| Mental symptoms | | |
| Development Well-being Assessment Interview[21] (DAWBA) | Major Depression | A sum of thirty-five depression-related question scores by self-report |
| | ADHD (child) | A sum of three ADHD-related question scores by self-report (i.e. "Child says teachers/teachers complain", " Child thinks self-hyperactive") |
| | ADHD (parent) | A sum of thirty ADHD-related question scores by parent-rated |

* indicates a reverse-coded preprocessing.

Table SM3. Socioeconomic, cognition and mental health assessments in UKB

| Variables | Field ID | Descriptions |
|---|---|---|
| Socioeconomics | | |
| Household income | 738 | Average total household income before tax, discretized by 18k, 40k, 52k and 100k |
| Indices of Multiple Deprivation (IMD) income score | 26411 (England)<br>26428 (Scotland)<br>26418 (Wales) | Indices of Multiple Deprivation come from a UK government qualitative study of deprived areas in British local councils. The study is conducted separately in England, Scotland and Wales with different components. In general, IMD was consisted of scores from crime, education, employment, health, housing, income, access to services and physical environment. |
| IMD employment score | 26412 (England)<br>26429 (Scotland)<br>26419 (Wales) | |
| IMD education score | 26414 (England) | |

| | | |
|---|---|---|
| | 26431 (Scotland)<br>26421 (Wales) | |
| Job physical | 816 | Job involves heavy manual or physical work |
| Neurocognition | | |
| Numeric memory test | 4282 | The participant was shown a 2-digit number to remember. The number then disappeared and after a short while they were asked to enter the number onto the screen. The number became one digit longer each time they remembered correctly (up to a maximum of 12 digits). We used maximum digits remembered correctly to measure numeric short-term memory |
| Trail making test | 6348, 6350 | The participant was presented with sets of digits/letters in circles scattered around the screen and asked to click on them sequentially according to a specific algorithm. We used the average time to complete numeric (trail #1) and alphanumeric path (trail #2) to measure visual attention and task switching |
| Tower rearranging test | 21004 | The participant was presented with an illustration of three pegs (towers) on which three differently-coloured hoops had been placed. The were then asked to indicate how many moves it would take to re-arrange the hoops into another specific position. We used the number of puzzles correct to measure executive functioning |
| Intelligence | 20016 | A sum of the number of correct answers given to the thirteen fluid intelligence questions |
| Education | 6138 | Recoded as: 4=College or University degree; 3=A levels/AS levels, NVQ or HND or HNC, other professional qualifications or equivalent; 2=O levels/GCSEs, CSEs or equivalent; 1=None of the |

| | | |
|---|---|---|
| | | above. The highest qualification was used |
| Mental health | | |
| Depression | 41270 | Diagnosed as depressive episode (F32) or recurrent depressive disorder (F33) |
| Anxious | 41270 | Diagnosed as generalized anxiety disorder (F41.1) |
| Neuroticism score | 20127 | An externally derived summary score of neuroticism, based on 12 neurotic behavior domains |
| Mood swing | 1920 | "Does your mood often go up and down?" |
| Miserableness | 1930 | "Do you ever feel 'just miserable' for no reason?" |
| Irritability | 1940 | "Are you an irritable person?" |
| Sensitivity | 1950 | "Are your feelings easily hurt?" |
| Fed-up feelings | 1960 | "Do you often feel 'fed-up'?" |
| Nervous feelings | 1970 | "Would you call yourself a nervous person?" |
| Suffer from 'nerves' | 2010 | "Do you suffer from 'nerves'?" |
| Tense | 1990 | "Would you call yourself tense or 'highly strung'?" |
| Worrier feelings | 1980 | "Are you a worrier?" |
| Loneliness | 2020 | "Do you often feel lonely?" |
| Guilty feelings | 2030 | "Are you often troubled by feelings of guilt?" |
| Risk taking | 2040 | "Would you describe yourself as someone who takes risks?" |
